# A comprehensive bioinformatics analysis to identify potential prognostic biomarkers among TNFSF superfamily in breast cancer

**DOI:** 10.1101/2024.06.11.24308760

**Authors:** Abolfazl Moradi, Farinaz Vafadar Esfahani, Ali Mohammadian

**Author notes:** These authors contributed equally to this work (Abolfazl Moradi, Farinaz Vafadar Esfahani).

## Abstract

Breast cancer (BC) is one of most important mortality factors among women therefore to find important genes in BC can help early diagnosis, treatment or prevention. *TNFSF* or tumor necrosis factor Superfamily have an important role in various cancers. In BC, some of studies have found dual roles for these genes. In this research, we conducted a comprehensive and detailed bioinformatics study on this family. UALCAN, TNMplot, UCSC Xena, GEPIA, The Human Protein Atlas, Kaplan–Meier plotter, bc-GenExMiner, cBioPortal, STRING, GeneMANIA, Enrichr, TIMER and shinyDepMap were used for analysis. We found that these genes play their role through the immune system and the high expression of eight *FASLG, LTB, TNF, TNFSF8, TNFSF10, TNFSF11, TNFSF12, TNFSF13* genes were positively associated with OS and RFS. Overall, our data showed that these genes can be considered as prognostic biomarkers. Further, our results suggest that this family has anti-tumor activity.

## 1. Introduction

Globally, Breast cancer (BC) is the most commonly diagnosed cancer with approximately 2.3 million new cases in 2020, accounting for 11.7% of all cancer cases(1). BC is an uncontrolled proliferation of breast cells, beginning in different areas of the breast and leading to malignancy(2). Based on receptor status and the gene expression pattern, BC is divided into four molecular subtypes including, Luminal A, Luminal B, human epidermal growth factor 2 (HER2), and Triple-negative breast cancer (TNBC).

Immunotherapy is one of the most exciting treatment strategies for breast cancer(3). To date, the US Food and Drug Administration has approved several immune-targeted therapies(4,5). Immune checkpoint inhibitors (ICIs), including inhibitors of cytotoxic T lymphocyte-associated antigen-4 (CTLA-4), programmed death-1 (PD1), and programmed death ligand-1 (PD-L1) have demonstrated a positive effect in the treatment of some cancers(6).

Although the methods of treatment and early detection of breast cancer have progressed, the prognosis of some patients remains poor due to metastasis and resistance to chemotherapy. Therefore, better therapeutic and prognostic biomarkers should be identified in breast cancer(7,8).

Tumor necrosis factor superfamily (TNFSF) has 19 members. The members of this family are expressed by immune cells and regulate the immune response and inflammation, proliferation, differentiation and apoptosis(9,10). During tumor development, inflammation in the tumor microenvironment (TME) causes tumor initiation, promotion, and progression(11).

In 1975 Carols discovered that *TNF* can inhibit cancer(12). Also, members of the TNFSF superfamily show pro-inflammatory activities by activating the NF-κB signaling pathway. Apart from these, it has been seen that they can activate apoptosis pathways and cause cell death. For example, some studies show that TNFSFs are tumor suppressor but others demonstrate they promote cancer(13).

*APRIL* (*TNFSF13*) has been found to be expressed in breast cancer cells, and Silencing experiments decreased cell proliferation, suggesting that *APRIL* is a critical factor for breast tumor growth(14).

However *TNFSF10, TNFSF14* and *TNFSF15* directly play a role in apoptosis(15). Some members of this family have been identified in lung cancer and kidney cancer(16,17).

According to different studies, this family have variety of roles(13). Therefore, we decided to investigate the role and activity of TNFSF family members comprehensively and more precisely in breast cancer.

## 2. Results

### 2.1 The mRNA expression analysis of TNFSF family in BC patients

The mRNA expression levels of TNFSF between primary tumor and normal tissues in BC patients were assessed using UALCAN on TCGA data. The mRNA expression levels of, *FASLG, LTA, LTB, TNF, TNFSF4, TNFSF8, TNFSF13B, TNFSF13* were found to be elevated in primary tumors compared to normal specimens, while *EDA, TNFSF12* were significantly downregulated in tumor samples (Fig 1). Also, the results showed that *TNFSF10* has the highest level of expression in breast cancer (fig 2).

**Fig 1:**
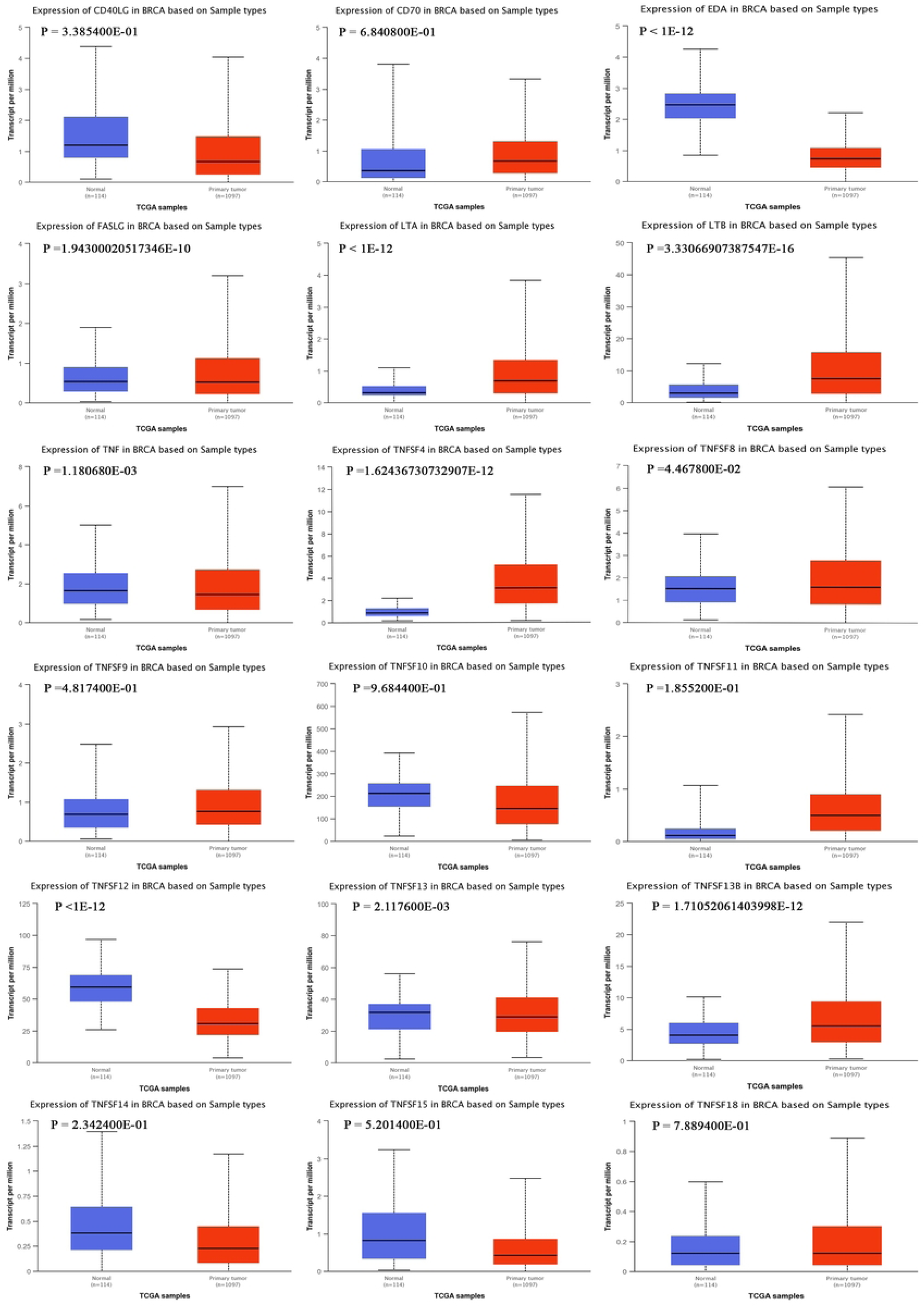
The difference in expression of TNFSFs in cancer and normal cells.

**Fig 2:**
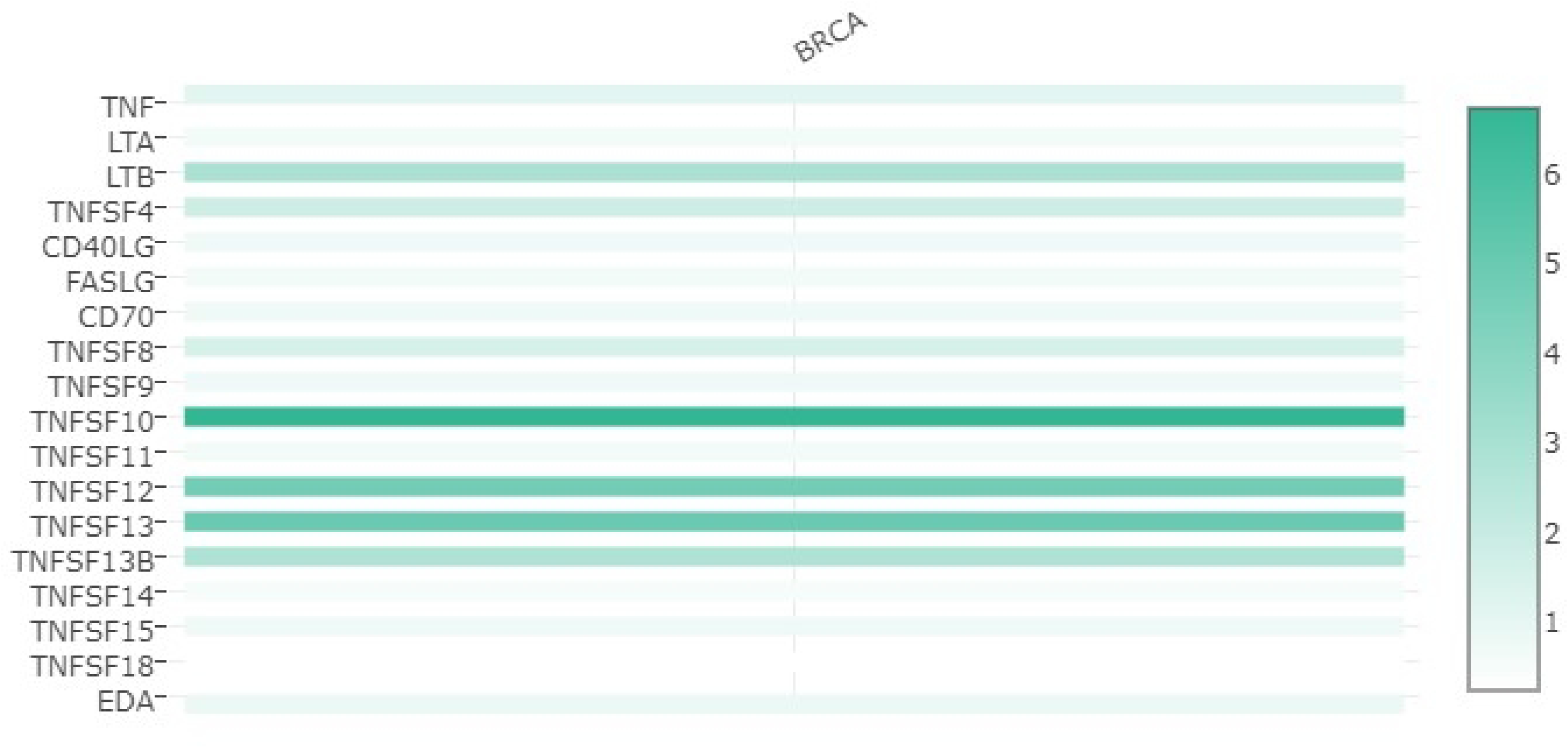
**The difference in expression of TNFSFs in breast cancer.**

### 2.2 Differential gene expression analysis in Tumor, and Metastatic tissue

Using the TNMplot site, the expression level of TNFSF superfamily genes was obtained, which genes EDA, CD40LG, LTA, CD70, LTB, *TNFSF4, TNFSF10, TNFSF11, TNFSF12, TNFSF13B, TNFSF14, TNFSF15* had a significant expression difference (P.value < 0.05). The analysis of FASLG, TNF, *TNFSF8, TNFSF9, TNFSF13, TNFSF18* genes did not show significant expression differences (P.value > 0.05), (fig 3).

**Fig 3:**
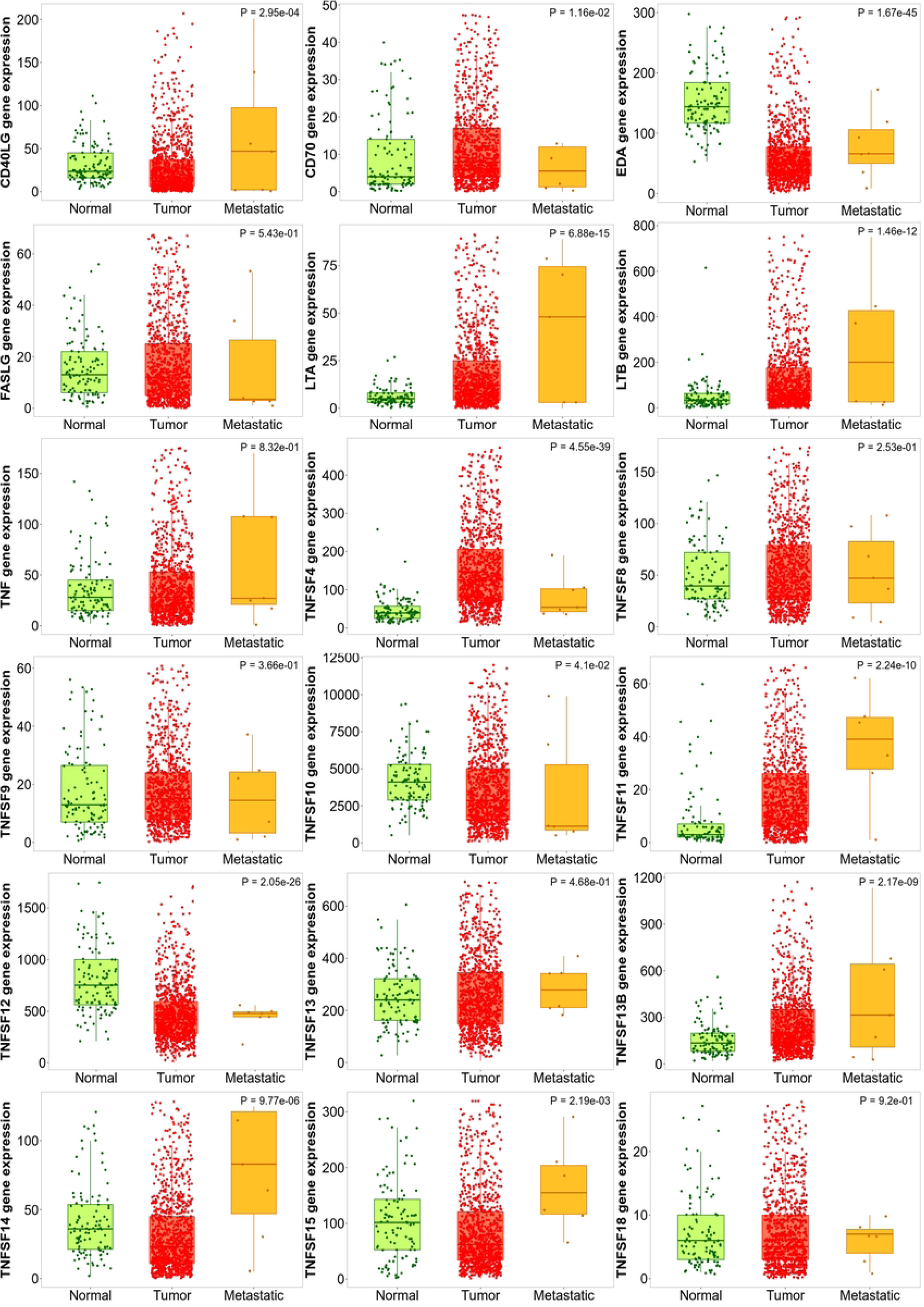
Differential gene expression analysis in Tumor, Normal, and Metastatic tissues.

### 2.3 TNFSFs Protein expression pattern analysis

In this study, we analyzed the expression pattern of proteins with the HPA database. *FASLG, CD40LG, LTB* proteins are not expressed in either normal or cancer tissues. Also, the expression of *LTA, TNF, TNFSF4* and *TNFSF12* proteins were not observed in normal tissue, but they were detected by antibodies in cancer tissue. Although *TNFSF10, TNFSF13* and *TNFSF13B* proteins were detected in normal tissue, they were present at higher levels in cancer tissue, and *TNFSF11*, *TNFSF15* proteins in both normal tissue and cancer tissue did not show difference in expression (fig 4).

**Fig 4:**
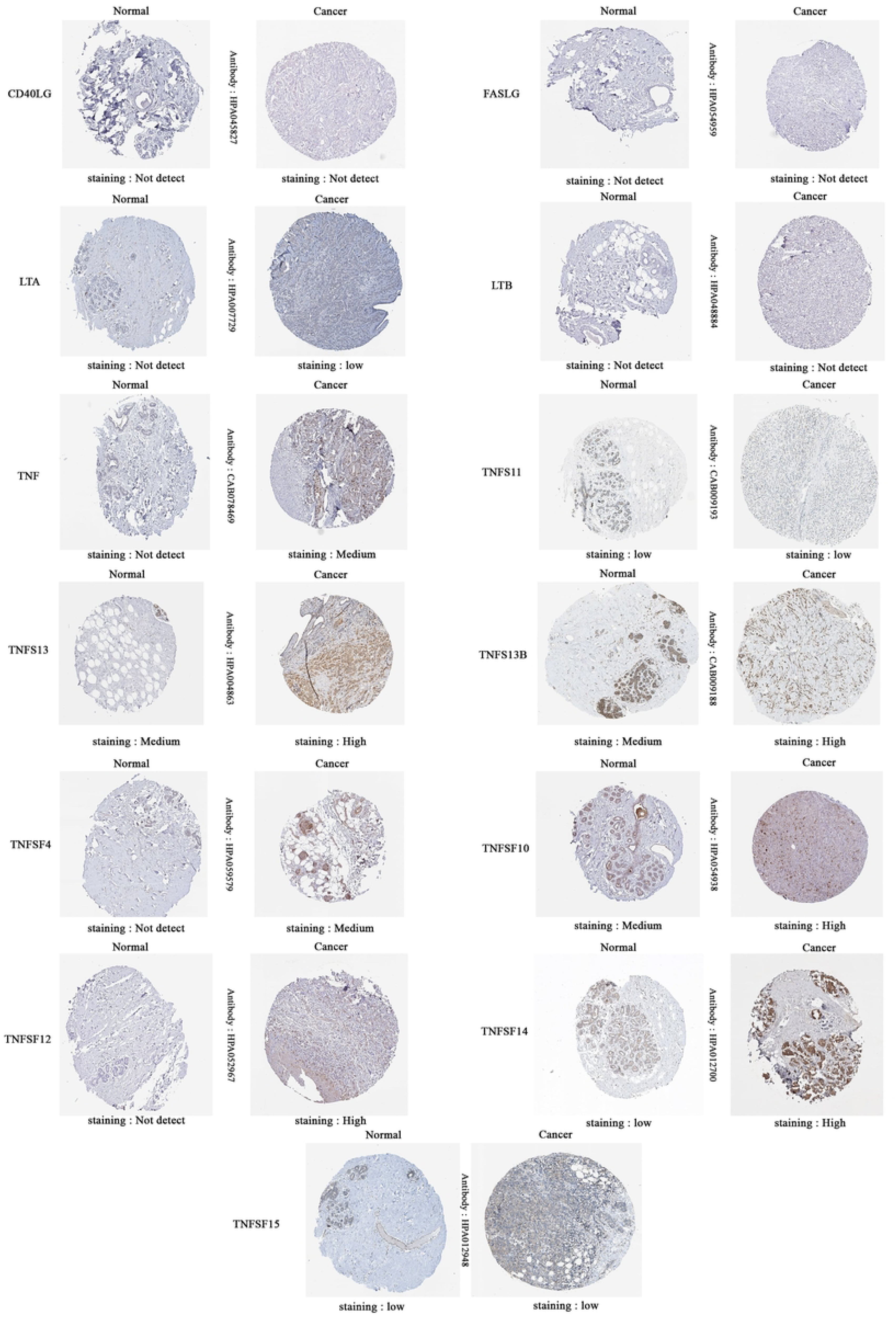
the expression pattern of proteins with the HPA database.

### 2.4 Association of TNFSFs mRNA levels with clinicopathological features in BC patients

By examining the results obtained from bC-genexminer (SBR), we found that the expression of *EDA*, *TNFSF10, TNFSF11, TNFSF12 and TNFSF13* decreased with tumor progression and the gene *TNFSF18* remained unchanged, while other family members showed an increase in expression with tumor progression (P < 0.05) (fig 5). The relationship between the expression TNFSFs and age showed that in younger patients (age≤51), *TNFSF11 and TNFSF15* had an increased expression, while in older patients (age>51) *TNFSF10 and TNFSF13* an increase in expression was observed (P < 0.05). In positive lymph nodes, the expression level of *ADE* was higher, but in negative lymph nodes, the expression level of *LTB*, *TNF, TNFSF14* was higher (P < 0.05). By comparing the results of ER+/ER- and PR+/PR-, in ER-/PR-the expression of *CD70, FASLG, LTA, LTB, TNF, TNFSF9, TNFSF13B, TNFSF14, TNFSF15* and in PR+/ER+, *TNFSF4, TNFSF10, TNFSF12, TNFSF13* were higher. Among common ER+/ER- and PR+/PR- genes, *CD40LG* showed lower expression in ER+, while higher expression was observed for this gene in PR+. *TNFSF8* was also higher expressed only in PR+ group(P < 0.05). In HER2, only the expression of *EDA, TNF, TNFSF12, TNFSF18, TNFSF4* and *TNFSF10* were significant (P < 0.05). The results indicate that in HER2 ^-^ the expression of *EDA, TNF, TNFSF12, TNFSF18* are higher than HER2^+^, but the *TNFSF4* and *TNFSF10* are less expressed. Similar results were observed in TNBC and Basal-like subtypes. Both models have a significant association with higher expression *CD40LG, CD70, FASLG, LTA, LTB, TNF, TNFSF9, TNFSF13B, TNFSF14, TNFSF15* and lower expression *TNFSF4, TNFSF10, TNFSF12, TNFSF13* (P < 0.05). All the results are presented in the table 1.

**Figure 5:**
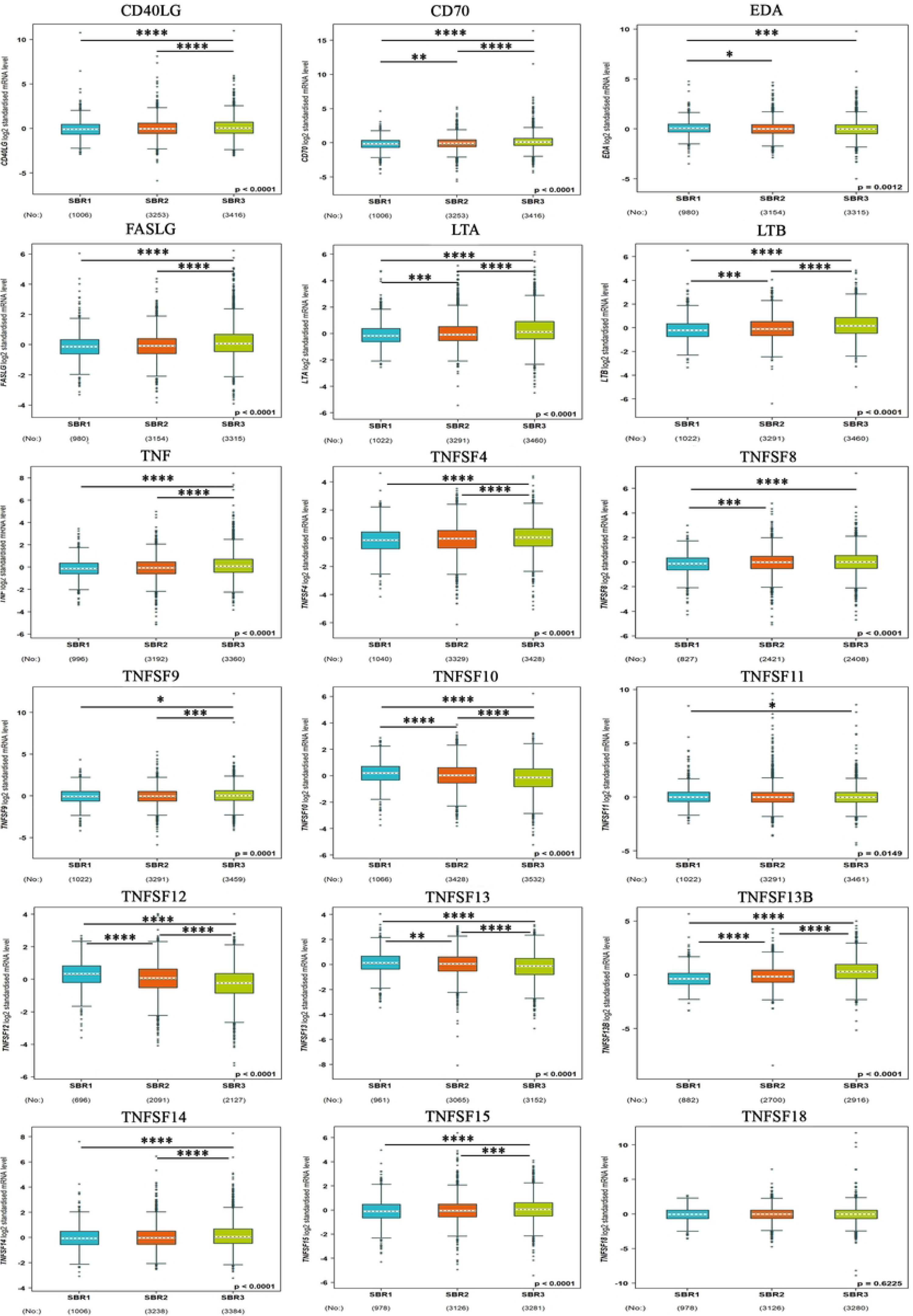
Association of TNFSFs mRNA levels with clinicopathological features in BC patients, **** = P < 0.0001, *** = P < 0.001, ** = P < 0.01, * = P < 0.05.

**Table 1:** Association of TNFSFs mRNA levels with clinicopathological features in BC patients.

| Criteria |  | Age |  | Nodal status |  | ER(IHC) |  | PR(IHC) |  | HER2(IHC) |  |  | TNBC | Basal-like BC |  |
| --- | --- | --- | --- | --- | --- | --- | --- | --- | --- | --- | --- | --- | --- | --- | --- |
|  |  | ≤51 | >51 | (-) | (+) | (-) | (+) | (-) | (+) | (-) | (+) | Not | TNBC | Not | Basal-like |
| No. |  | 267 | 476 | 332 | 358 | 187 | 530 | 243 | 470 | 396 | 109 | 578 | 87 | 605 | 136 |
| CD40LG | mRna | - | - | - | - | ↑ | - | - | ↑ | - | - | - | ↑ | - | ↑ |
|  | P-value | 0.0643 |  | 0.9995 |  | 0.0019 |  | 0.0455 |  | 0.2442 |  | 0.0123 |  | 0.0102 |  |
| CD70 | mRna | - | - | - | - | ↑ | - | ↑ | - | - | - | - | ↑ | - | ↑ |
|  | P-value | 0.5767 |  | 0.061 |  | 0.0001 |  | 0.0001 |  | 0.3145 |  | 0.0001 |  | 0.0001 |  |
| EDA | mRna | - | - | - | ↑ | - | - | - | - | ↑ | - | - | - | - | - |
|  | P-value | 0.8397 |  | 0.0458 |  | 0.1625 |  | 0.6614 |  | 0.0001 |  | 0.4528 |  | 0.8725 |  |
| FASLG | mRna | - | - | - | - | ↑ | - | ↑ | - | - | - | - | ↑ | - | ↑ |
|  | P-value | 0.8256 |  | 0.2115 |  | 0.0001 |  | 0.0137 |  | 0.7380 |  | 0.0010 |  | 0.0002 |  |
| LTA | mRna | - | - | - | - | ↑ | - | ↑ | - | - | - | - | ↑ | - | ↑ |
|  | P-value | 0.3227 |  | 0.0713 |  | 0.0001 |  | 0.0001 |  | 0.5161 |  | 0.0001 |  | 0.0001 |  |
| LTB | mRna | - | - | ↑ | - | ↑ | - | ↑ | - | - | - | - | ↑ | - | ↑ |
|  | P-value | 0.1399 |  | 0.0476 |  | 0.0001 |  | 0.0001 |  | 0.1034 |  | 0.0001 |  | 0.0001 |  |
| TNF | mRna | - | - | ↑ | - | ↑ | - | ↑ | - | ↑ | - | - | ↑ | - | ↑ |
|  | P-value | 0.3435 |  | 0.0011 |  | 0.0001 |  | 0.0001 |  | 0.0034 |  | 0.0001 |  | 0.0001 |  |
| TNFSF4 | mRna | - | - | - | - | - | ↑ | - | ↑ | - | ↑ | ↑ | - | ↑ | - |
|  | P-value | 0.8480 |  | 0.9974 |  | 0.0011 |  | 0.0001 |  | 0.0008 |  | 0.0006 |  | 0.0001 |  |
| TNFSF8 | mRna | - | - | - | - | - | - | - | ↑ | - | - | - | - | - | - |
|  | P-value | 0.9179 |  | 0.7221 |  | 0.3911 |  | 0.0240 |  | 0.2112 |  | 0.5226 |  | 0.2062 |  |
| TNFSF9 | mRna | - | - | - | - | ↑ | - | ↑ | - | - | - | - | ↑ | - | ↑ |
|  | P-value | 0.6572 |  | 0.7787 |  | 0.0001 |  | 0.0001 |  | 0.0597 |  | 0.0003 |  | 0.0001 |  |
| TNFSF10 | mRna | - | ↑ | - | - | - | ↑ | - | ↑ | - | ↑ | ↑ | - | ↑ | - |
|  | P-value | 0.0032 |  | 0.7554 |  | 0.0001 |  | 0.0001 |  | 0.0075 |  | 0.0007 |  | 0.0001 |  |
| TNFSF11 | mRna | ↑ | - | - | - | - | - | - | - | - | - | - | - | - | - |
|  | P-value | 0.0001 |  | 0.1164 |  | 0.6868 |  | 0.1989 |  | 0.3689 |  | 0.9446 |  | 0.3443 |  |
| TNFSF12 | mRna | - | - | - | - | - | ↑ | - | ↑ | ↑ | - | ↑ | - | ↑ | - |
|  | P-value | 0.256<br>2 |  | 0.17<br>27 |  | 0.0001 |  | 0.0001 |  | 0.000<br>1 |  | 0.0003 |  | 0.0001 |  |
| TNFSF13 | mRna | - | ↑ | - | - | - | ↑ | - | ↑ | - | - | ↑ | - | ↑ | - |
|  | P-value | 0.004<br>2 |  | 0.21<br>11 |  | 0.0001 |  | 0.0001 |  | 0.127<br>5 |  | 0.0001 |  | 0.0001 |  |
| TNFSF13<br>B | mRna | - | - | - | - | ↑ | - | ↑ | - | - | - | - | ↑ | - | ↑ |
|  | P-value | 0.913<br>1 |  | 0.15<br>23 |  | 0.0001 |  | 0.0006 |  | 0.664<br>1 |  | 0.0001 |  | 0.0001 |  |
| TNFSF14 | mRna | - | - | ↑ | - | ↑ | - | ↑ | - | - | - | - | ↑ | - | ↑ |
|  | P-value | 0.460<br>3 |  | 0.02<br>36 |  | 0.0001 |  | 0.0001 |  | 0.209<br>2 |  | 0.0001 |  | 0.0001 |  |
| TNFSF15 | mRna | ↑ | - | - | - | ↑ | - | ↑ | - | - | - | - | ↑ | - | ↑ |
|  | P-value | 0.001<br>0 |  | 0.15<br>35 |  | 0.0001 |  | 0.0001 |  | 0.601<br>5 |  | 0.0001 |  | 0.0001 |  |
| TNFSF18 | mRna | - | - | - | - | - | - | - | - | ↑ | - | - | - | - | - |
|  | P-value | 0.077<br>9 |  | 0.50<br>43 |  | 0.1489 |  | 0.8882 |  | 0.035<br>1 |  | 0.4528 |  | 0.2337 |  |

### 2.5 Genomic alterations and GO enrichment analysis of TNFSF members in BC patients

The data extracted from cBioprotal database and 1180 cancer samples (TCGA, Firehose Legacy) showed that in breast cancer genes *TNF* (4%), *LTA* (4%), *LTB* (3%), *TNFSF4* (12%), *CD40LG* (3%), *FASLG* (11%), *CD70* (3%), *TNFSF8* (3%), *TNSF9* (3%), *TNFSF10* (7%), *TNFSF11* (4%), *TNFSF12* (5%), *TNFSF13* (1%), *TNFSF13B* (4%), *TNFSF14* (3%), *TNFSF15* (3%), *TNFSF18* (11%), *EDA* (4%) were altered (fig 6A). In addition, by using the STRING, GeneMANIA database and Cytoscape software, the protein-protein interaction network and Interaction at the gene level of the TNFSF superfamily members and the top 50 frequently altered neighbor genes co-expressed genes were mapped (fig 6 D,F). Genomic alterations of the top 50 frequently co-altered genes with TNFSF Superfamily members in BC patients are presented in S 1 Table. In the next step, the function and pathway of the respiratory superfamily proteins and the altered neighbor genes were investigated. According to the results obtained from Enrichr, the most important function of members of this family with altered neighbor genes is tumor necrosis factor receptor superfamily binding (fig 6C). In KEGG pathway analysis, we found that TNFSFs and their neighbor genes were most commonly enriched in Cytokine-cytokine rreceptor interaction (Fig 6B).

**Fig 6:**
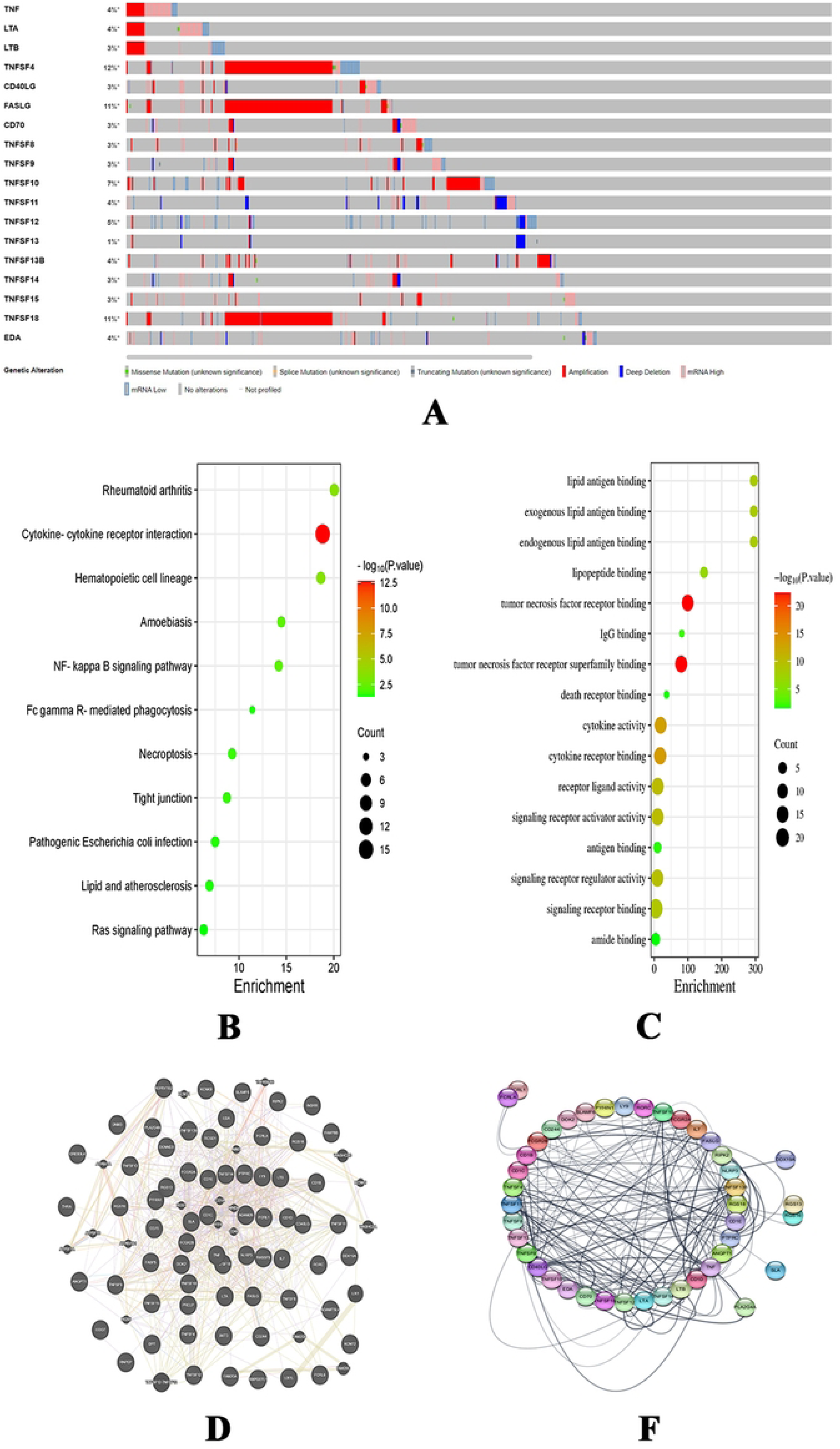
Genomic alterations and GO enrichment analysis of TNFSF members in BC patients. (A): Genomic alteration of TNFSFs, (B): KEGG pathway analysis, (C): function analysis, (D): Gene Interactions analysis, (F): the protein-protein interaction network.

### 2.6 Prediction of transcription factors (TFs) and miRNA related to TNFSF superfamily

Transcription factors and microarrays regulating TNFSF genes were identified using ChEA and miRTarBase databases, and the obtained results are reported in tables 2 and 3.

**Table 2:**
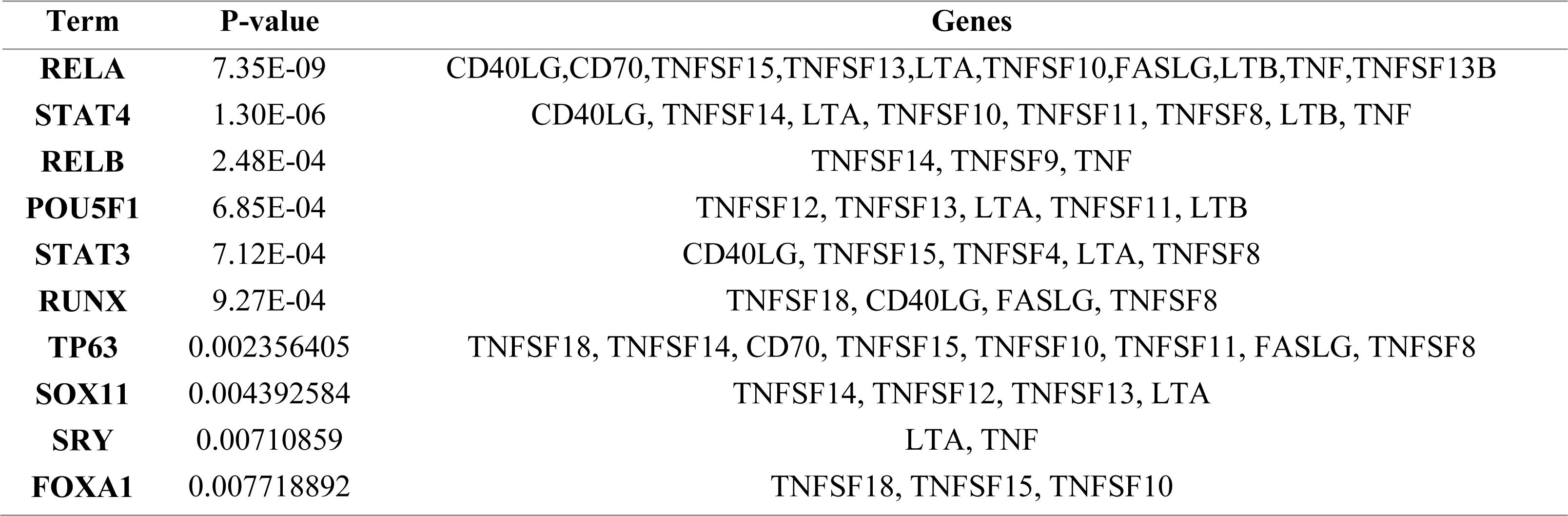
transcription factors (TFs) related to TNFSF superfamily.

**Table 3:** miRNA related to TNFSF superfamily.

| Term | P-value | Genes |
| --- | --- | --- |
| **hsa-miR-34a-5p** | 3.80E-04 | CD40LG, CD70, LTA, FASLG, TNF |
| **hsa-miR-6510-5p** | 0.004722 | TNFSF9, TNFSF8 |
| **hsa-miR-4480** | 0.005553 | TNFSF14, TNFSF9 |
| **hsa-miR-3619-5p** | 0.007901 | TNFSF15, TNFSF9 |
| **hsa-miR-761** | 0.008106 | TNFSF15, TNFSF9 |
| **mmu-miR-296-3p** | 0.01075 | TNF |
| **hsa-miR-9500** | 0.013963 | CD40LG, TNFSF8 |
| **hsa-miR-202-3p** | 0.014493 | TNFSF9, TNFSF13B |
| **hsa-miR-214-3p** | 0.014493 | TNFSF15, TNFSF9 |
| **mmu-miR-212-3p** | 0.016083 | TNFSF10 |

### 2.7 The prognostic value of TNFSF in patients with BC

The Kaplan–Meier curves revealed that among TNFSFs, high mRNA expression of *FASLG, LTB, TNF, TNFSF8, TNFSF10, TNFSF11, TNFSF12, TNFSF13* were significantly associated with better OS (P < 0.05) (fig 7).

**Fig 7:**
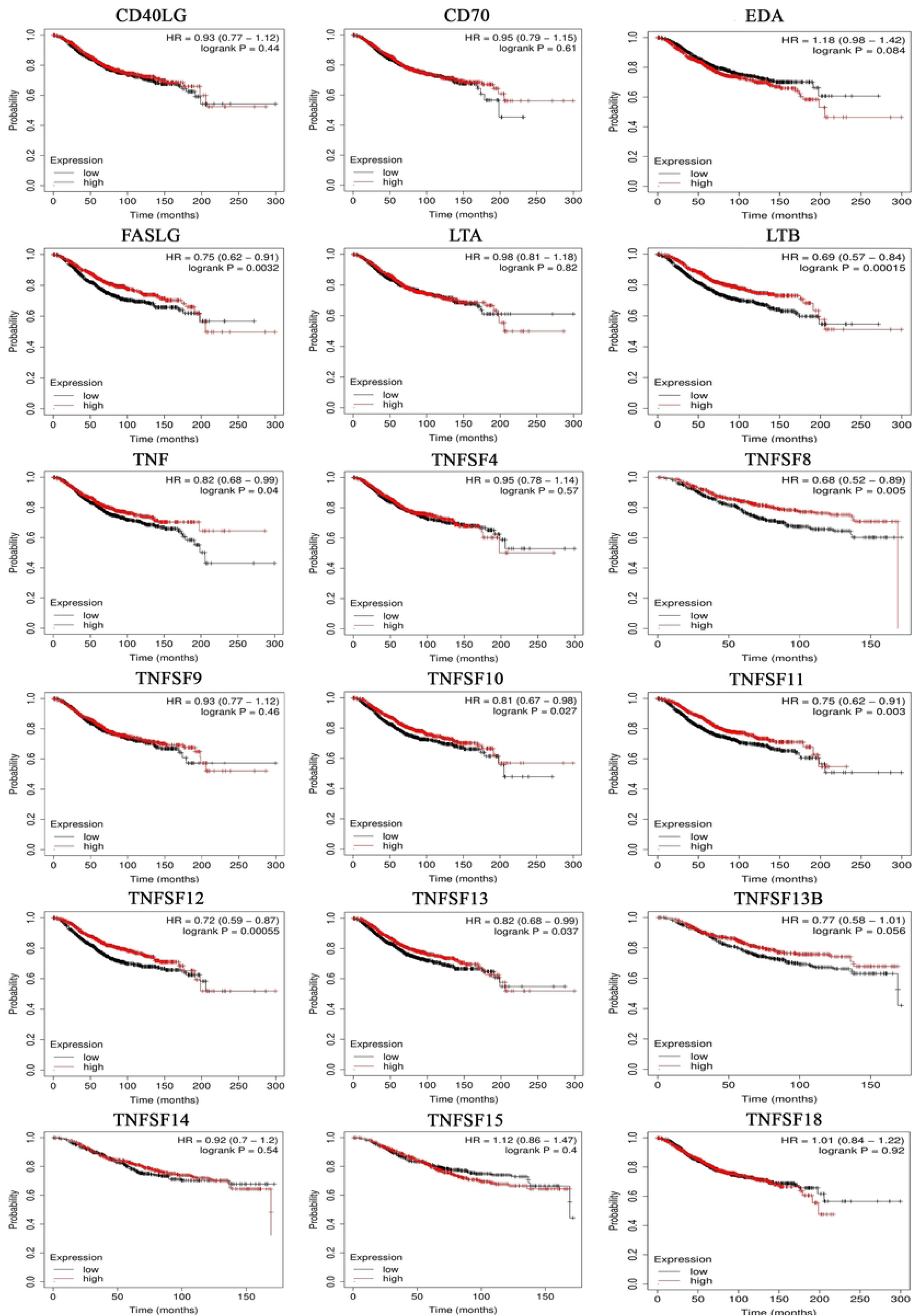
The prognostic value (OS) of TNFSF in patients with BC.

In addition, regarding RFS, BC patients with increased mRNA levels of *CD40LG, EDA, FASLG, LTA, LTB, TNF, TNFDF8, TNFDF9, TNFDF10, TNFDF11, TNFDF12, TNFDF13, TNFDF14, TNFDF15, TNFDF18* were significantly correlated with favorable RFS (P < 0.05) (fig 8). On the other hand, elevated expression of *TNFSF4, CD70, TNFSF13B* were remarkably correlated with unfavorable RFS (P < 0.05). curves of chemokines in which mRNA expression levels are significantly associated with OS and RFS (fig 8).

**Fig 8:**
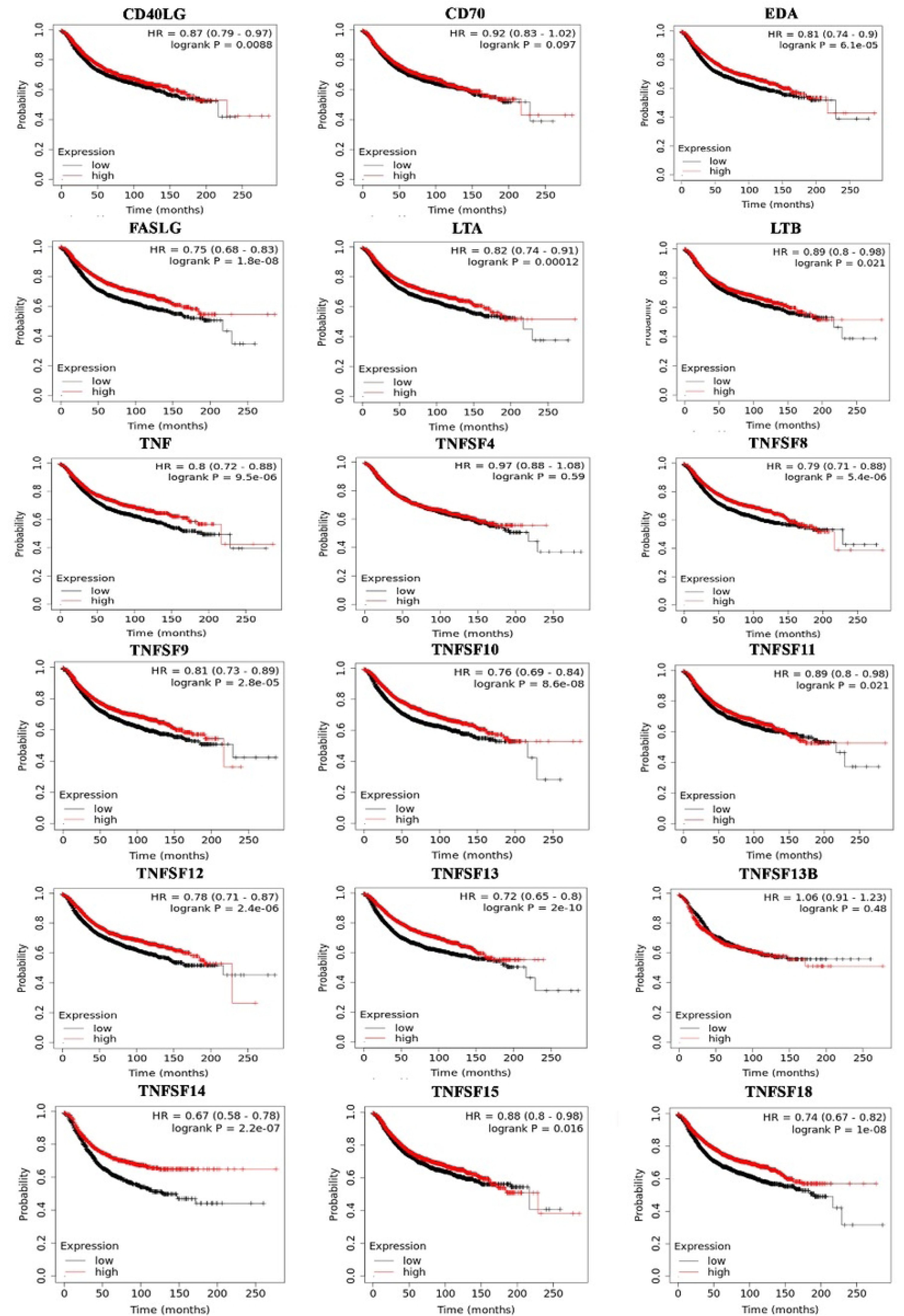
The prognostic value (RFS) of TNFSF in patients with BC.

Considering that *FASLG, LTB, TNF, TNFSF8, TNFSF10, TNFSF11, TNFSF12, TNFSF13* genes were correlated with favorable both OS and RFS, their correlation with OS in breast cancer subtypes (fig 9) and also in different grades (fig 10) was checked. The results showed that the high expression of *TNFSF11* gene in luminal A has a favorable correlation with OS (P < 0.05), but patients with luminal B and lower expression of *TNFSF11* had better survival. The results of *TNFSF8* analysis in breast cancer subgroup showed a significant correlation with luminal A, luminal B and Basal-Like, and the group of patients with higher expression had better OS (P < 0.05). For *TNFSF12*, only one significant correlation was observed in breast cancer subgroups, and its high expression showed a favorable correlation with OS in patients with HER2 (P < 0.05). Patients with lower expression of *LTB, TNFSF8, TNFSF10, TNFSF13, FASLG* genes all had shorter survival.

**Figure 9:**
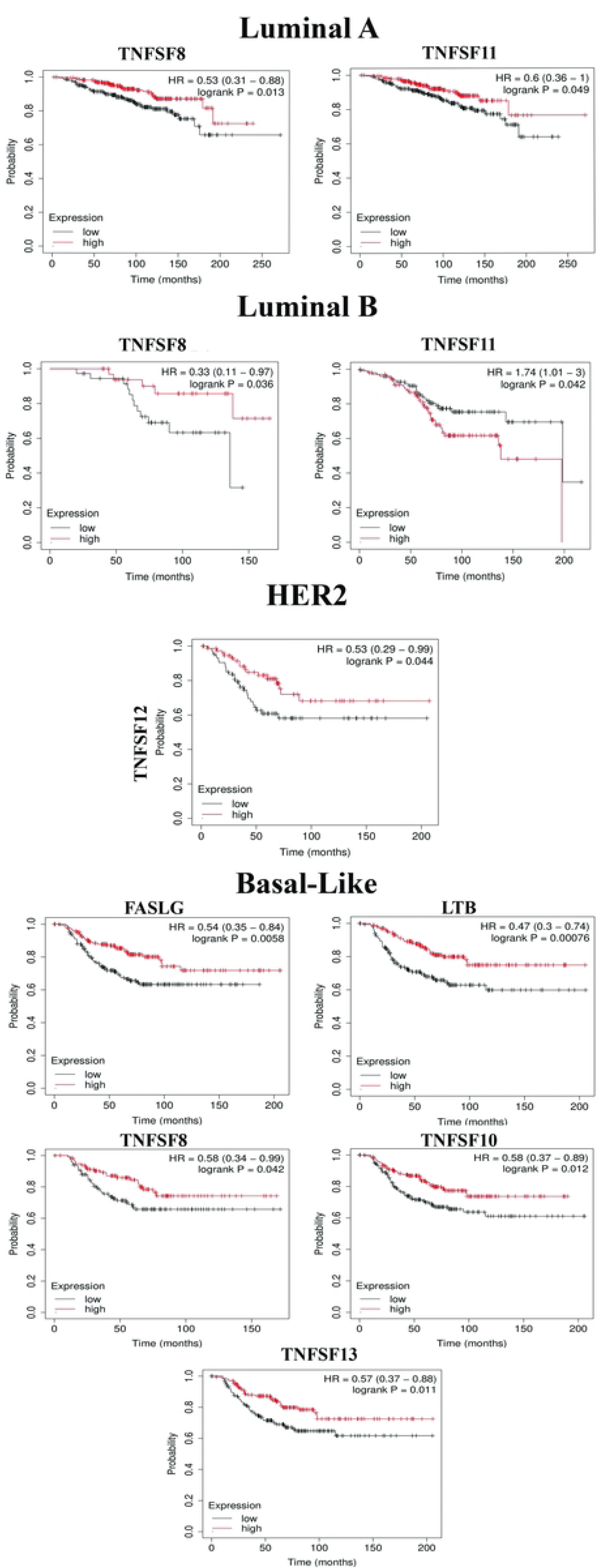
The prognostic value (OS) of TNFSF in sub-type of BC.

**Fig 10:**
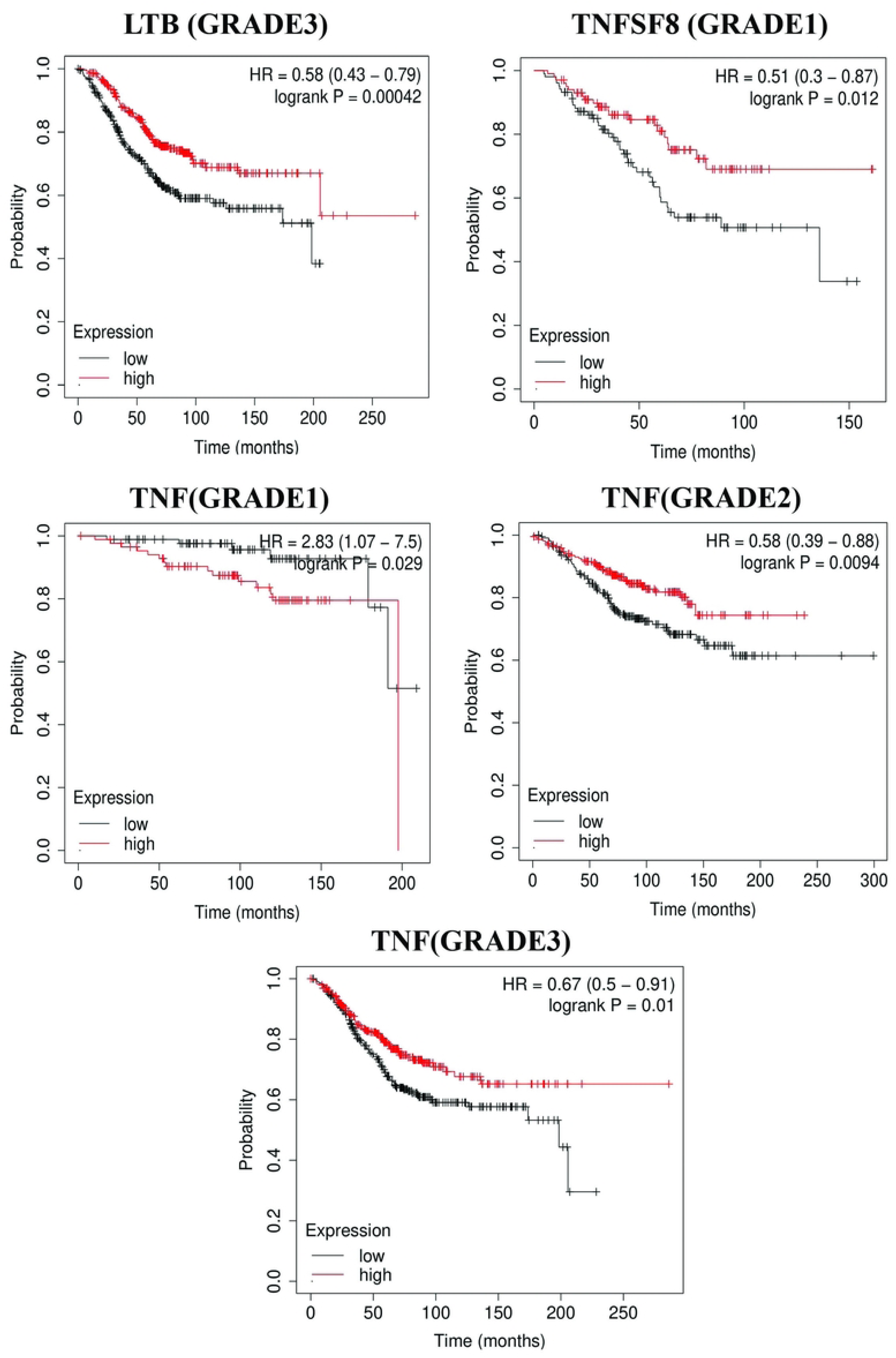
The prognostic value (OS) of TNFSF in grade of BC.

Survival analysis based on different grades of breast cancer showed that *LTB* only in grade 3, *TNFSF8* in grade 1 and *TNF* had a significant correlation with OS in all disease grade (P < 0.05).

Among these three genes, the lower expression of *TNF* in grade 1 had a favorable correlation with OS, but in grades 2 and 3 it is completely opposite.

### 2.8 The prognostic value of TNFSF in patients with metastases

Survival analysis and data extracted from UCSC Xena (Breast Cancer (Vijver 2002) didn’t show any significant correlation between the expression of TNFSF (except *TNFSF14*) in metastatic tissue and survival of patients. *TNFSF14* had a Pvalue less than 0.05. (Fig 11).

**Fig 11:**
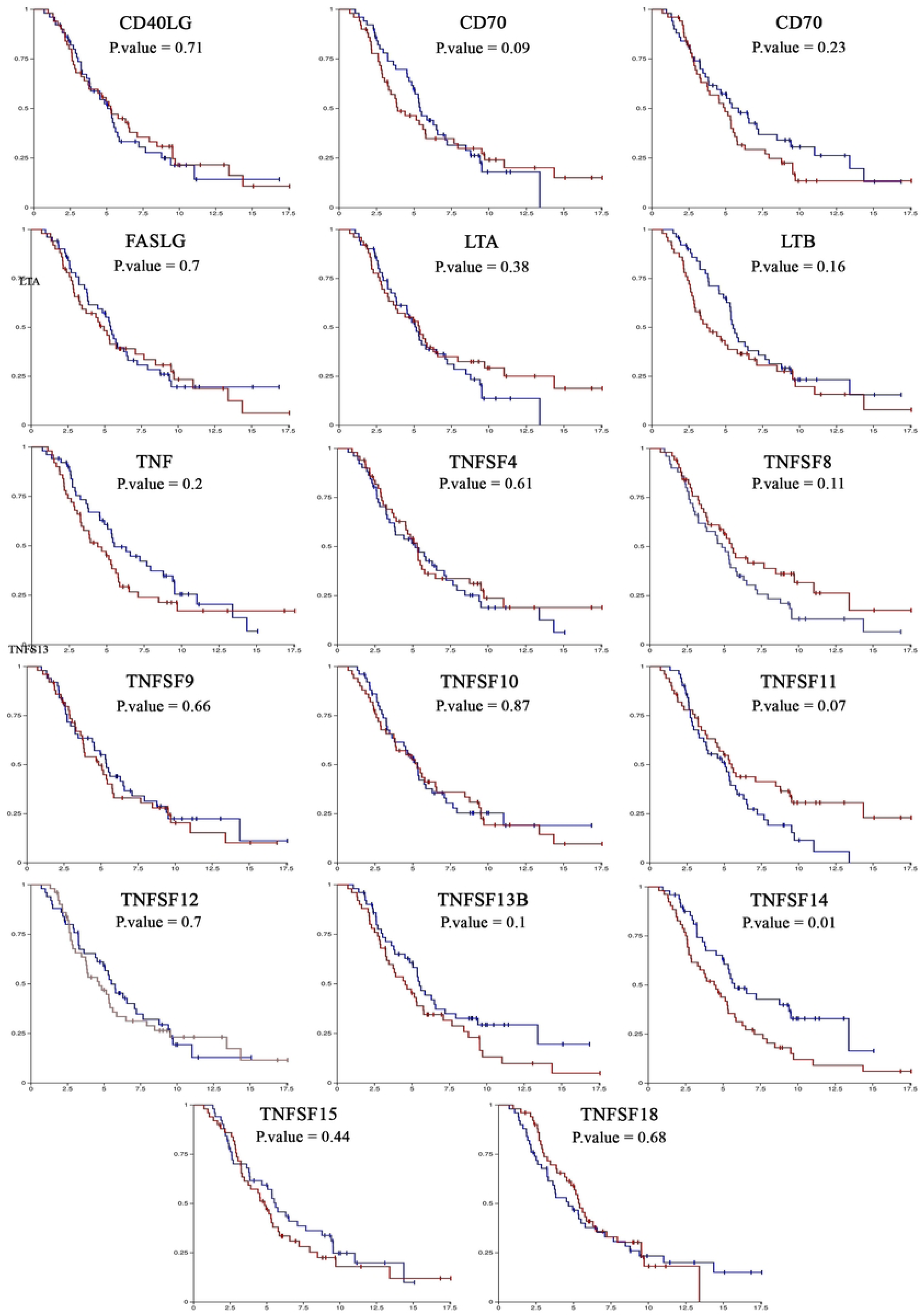
The prognostic value of TNFSF in patients with metastases.

### 2.9 Correlation between TNFSFs and immune cell infiltration in BC

The results obtained from the TIMER database prove that apart from *TNF* with macrophage infiltration (P.value = 0.591), EDA with B cell infiltration (P.value = 0.079), *TNFSF13* with CD8^+^ Tcell infiltration(P.value = 0.0601) and *TNFSF12* with B cell and CD8^+^ Tcell infiltration(P.value = 0.208) the expression of the genes TNFSFs are positively correlated with the infiltration of immune cells (B cell, CD8+ T cell, CD4+ T cell, macrophage cells, neutrophil cells, and dendritic cells) (all with P<0.05) (S 2 and 3 Figs). Also, the table 4 shows the expression of TNFSFs was mostly correlated with high infltration abundances of which of immune cells.

**Table 4::**
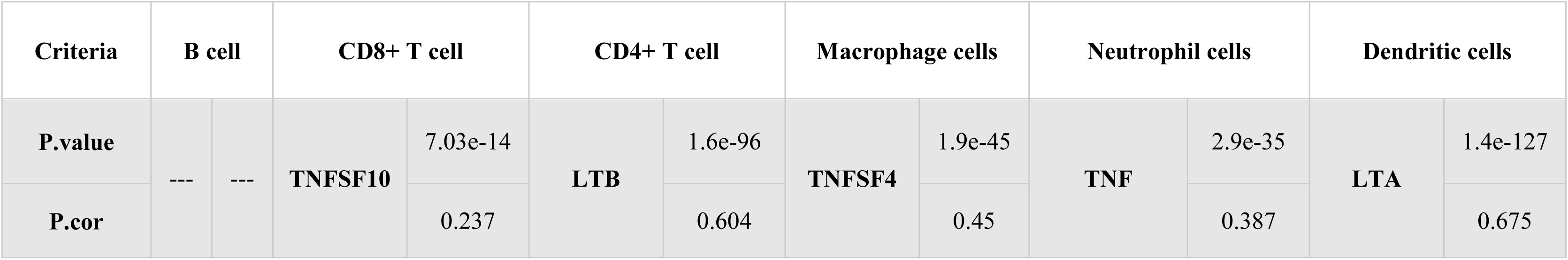
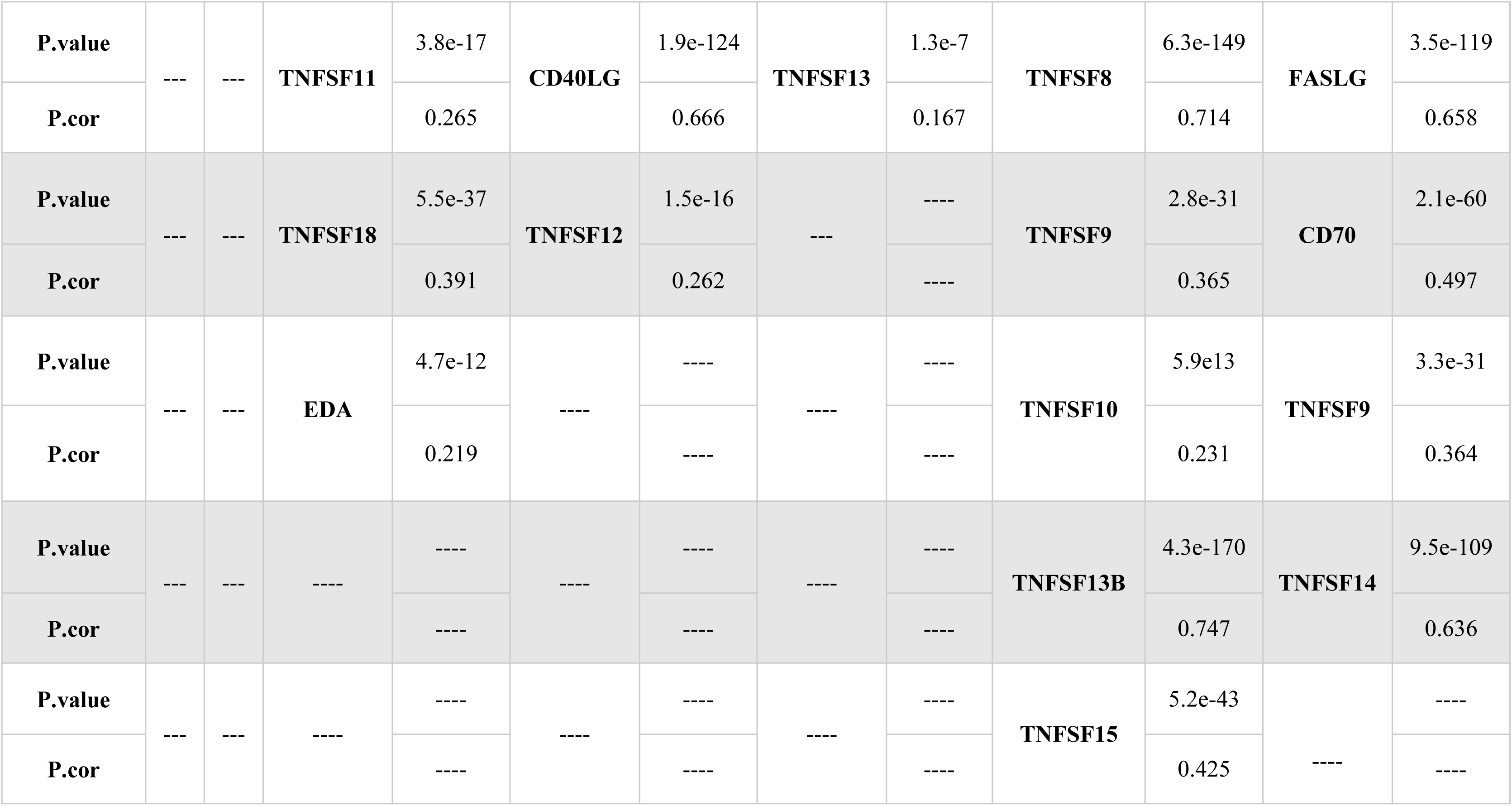
Correlation between TNFSFs and immune cell infiltration in BC.

### 2.10 Gene essentiality of TNFSF family genes

DepMap analysis confirmed none of TNFSFs weren’t essential for breast cancer, even these genes are not suitable targets for drugs. According to the obtained results and Fig 12, even the efficiency and selectivity factor have not reached the threshold of the software. Only in one case, *TNFS10*, more selectivity and efficiency were observed, but the analysis states that this gene is essential for kidney cancer than breast cancer.

**Fig 12:**
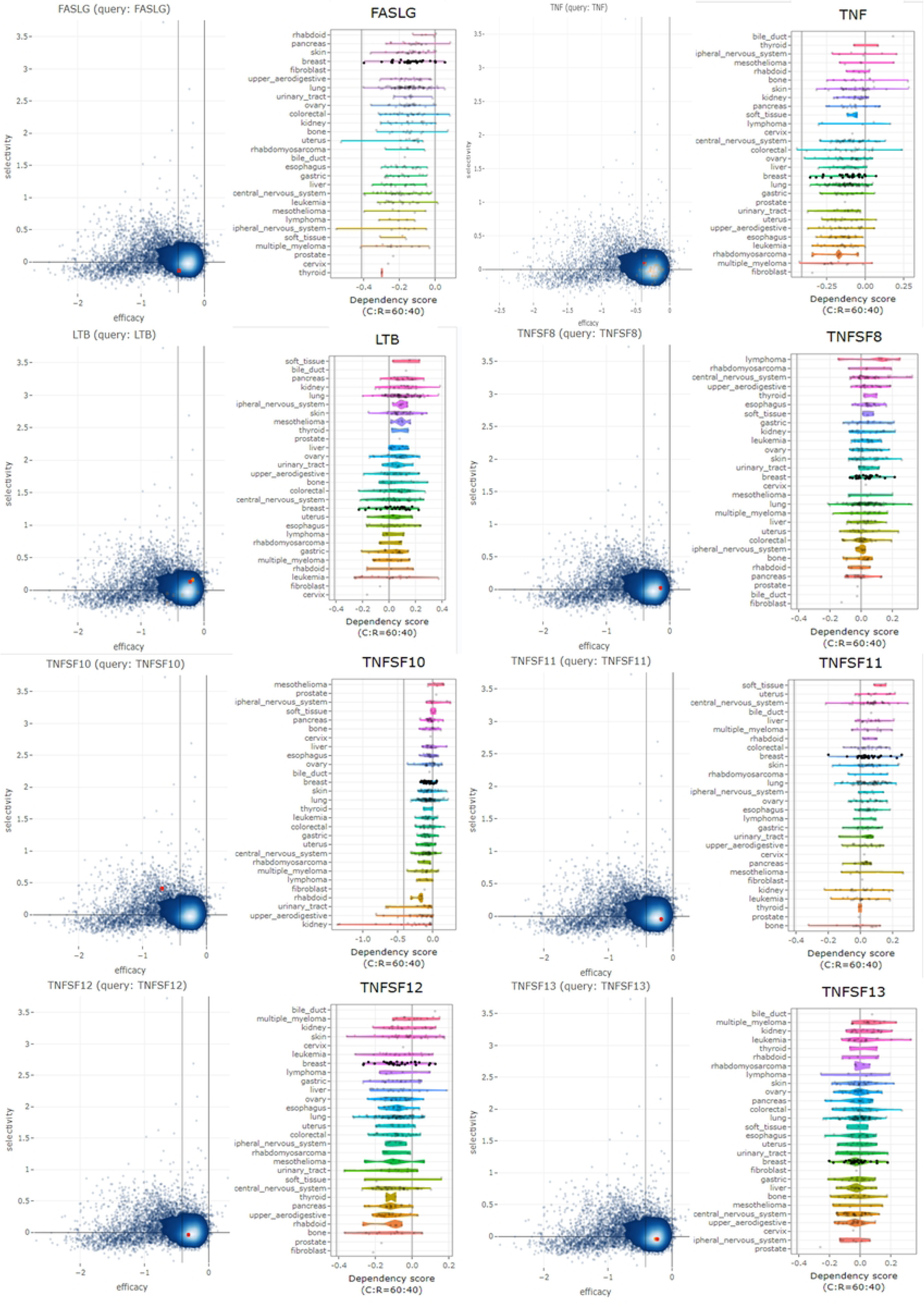
Gene essentiality of TNFSF family genes.

## 3. Discussion

Considerable data have clearly shown that the TNFSF family can promote cancer development, while in some cases the opposite result has been obtained. In the current study, we applied a bioinformatic approach to elucidate the prognostic values of the whole TNFSF family in BC.At first, the We found that 10 genes (*EDA, FASLG, LTA, LTB, TNF, TNFSF4, TNFSF8, TNFSF12, TNFSF13, TNFSF13B*) are differently expressed in TCGA breast cancer data. The expression of five these 10 genes were also assessed in different SBR stages. Five genes (*EDA, TNFSF10/11/12/13*), were found to decrease with tumor progression. *TNFSF18* expression was constant during tumor progression. The other genes showed increased expression. In addition, according to the obtained results from TNMplot, *CD40LG, LTA, CD70, LTB, TNFSF4, TNFSF10, TNFSF11, TNFSF12, TNFSF13B, TNFSF14, TNFSF15* genes were down-regulated in metastatic tissue relative to tumor cells. The rest of the genes did not show significant difference. The most genetic alternation among TNFSF family members was amplification.

Survival analysis and Kaplan-meier plotter confirmed prognostic value *of LTB, FASLG, TNF, TNFSF8/10/11/12/13.* Higher expression of these genes was associated with both longer overall survival and recurrence-free survival. Although *CD40LG, LTA, TNFSF9/14/15/18* genes had weak association with OS but high expression of those genes had favorable correlation with RFS. The prognostic value of the TNFSFs was significant just for *TNFSF14* in metastatic tissue that patients with lower expression showed better survival.

The results obtained from bc-GenExMiner v4.8 showed that TNF gene had a better correlation with nodal status^-^, PR^-^, ER^-^, HER2^-^, TNBC and Basal-like. In addition, higher expression of *TNF* was observed in TNBC, Basal-Like, nodal status^-^, PR^-^, ER^-^, HER2^-^. Our results confirmed that the expression of this gene increases with progression of cancer. Immunohistochemistry also shows an increase in expression in cancer cells. Better OS by increasing the expression of this gene states that *TNF* plays an important role in suppressing cancer or tumor, although recent researches have envisioned a dual role for this gene. In 1975, it was shown that *TNF* is a necrosis factor and destructs tumor cells indirect (18,19). It has been found that administration of *TNF* may induce apoptosis in malignant cells(19,20). In addition, it has the ability to cause inflammation(21) and tumor growth(22–24) but in certain cases it induces apoptosis. *TNF* performs different activities depending on the specific cell. For example, this gene stimulates the proliferation of *T47D*(25,26) and it has apoptotic(27–29) and anti-mitogenic(30,31) roles for *MCF7*. For this reason, we tried to investigate the expression level of this gene in metastatic tissue compared to tumor tissue and the correlation the expression of *TNF* with patient survival in metastatic tissues that our data didn’t show any significant difference in expression compared to tumor tissue and metastasis. Also, the expression of this gene had no significant correlation with OS in metastatic tissues.

According to the results extracted from the SBR analysis, the expression of *TNF* increases with the progress of the tumor, therefore, survival analysis was conducted in different grades and subtypes of breast cancer to understand function of *TNF*. We found out the expression of *TNF* didn’t had any correlation with OS in none of subtype of breast cancer but there was a correlation with OS in grade 1 where the patients with lower expression had better OS while everything was opposite in other grads. It is good to know, the result of DepMap stats that *TNF* is not an essential gene for breast cancer and even It has no medicinal value in different cancer so it seems that *TNF* exerts its effect on breast cancer through immune cells.

Tumor cells in the tumor microenvironment (TME) can directly invade the surrounding tissues or metastasize through blood and lymphatic vessels, and infiltrated cells can release cytokines, cytokine receptors, and other factors that directly or indirectly inhibit and promote tumor progression. Tumor cells progress, induce the immune response(32). In this study, we understood that *TNF* is related to the infiltration of immune cells into the tumor microenvironment, especially neutrophils (P.val = 2.9e-35, P.cor = 0.387). It has even been stated that this influence can contribute to the development of cancer and reduce survival of patients(33,34) so it should be studied how does cancer cells use immune cells because DepMap analysis introduces TNF as an unnecessary gene for breast cancer.

*FASLG* starts apoptosis by binding to their receptor (FAS) (35) as a result *FASLG* consider as toumor suppressor in different cancers by initiating and inducing apoptosis signaling(35–37). Although the increased expression of *FASLG* was not observed in the immunohistochemical data, these results are consistent with the data obtained from UALCAN and survival analysis. In addition, Clinical-pathological features and SBR results showed increased expression of *FASLG* and also, we found out it is correlated with TNBC, and survival of Basal-Like patients. Survival analysis confirmed the prognostic value of this gene and showed increased expression of *FASLG* are correlated with favorable OS and RFS. However, experiments have proven that this gene is involved in development of cancer. *FASLG* gene expression is increased in breast cancer. Also, this increasing causes apoptosis in T cells carrying Fas, which provides an advantage for cancer cells (38,39). Therefore, the survival analysis in different cancer grades and metastatic tissues was investigated. No correlation was found. Moreover, no significant expression difference was observed in metastatic tissues compared to tumor and normal tissues. We came to the conclusion that *FASLG* gene expression is related to infiltration of immune cells, especially dendritic cells (P.val = 3.5e-119, P.cor = 0.658). Dendritic cells infiltrating the tumor microenvironment have a good correlation with PFS and these cells initiate the immune response in breast cancer(40). Other studies confirm this result, for example, Coventry and Morton observed greater survival in breast cancer patients with a higher density of dendritic cells(41). It turns out this gene also plays its role in breast cancer through the immune system because DepMap does not consider this gene necessary for breast cancer.

*TNFSF10*, the other member of TNFSFs which is a Apoptosis inducer factor in cancer cells(42,43). *TNFSF10* can destroy the cell components and cause the process of apoptosis by binding to its receptor and by recruiting the adapter molecule FADD and activating caspases. There are some evidence that show the anti-tumor role this gene in cancer cells and microenvironment in addition not only this gene regulate apoptosis but also it has on effect on proliferation of immune cells and It stimulates the growth of M2 macrophages(44). There weren’t any significant changes in UALCAN data but increased expression of TNFSF10 was observed in obtained data from Human Protein Atlas database and survival analysis expresses that higher level of *TNFSF10* mRna has better correlation with survival of patients. We observed a significant correlation between expression of this gene and infiltration of immune cells, especially neutrophils (P.val =5.9e13, P.cor = 0.231) in TIMER result. According to explanations for TNF, Neutrophils has a role in the progression of cancer in the tumor microenvironment and it seems *TNFSF10* do its tasks in breast cancer through the immune system because DepMap does not consider this gene necessary for breast cancer. We found that as the tumor progresses, the expression of this gene decreases in cancer cells. Our results are the same as those obtained from the study on uveal melanoma. The expression of this gene decreases with the progression of metastasis and has been introduced as a tumor suppressor gene(45). These results are completely consistent with the data extracted from TNMplot. We observed that metastatic tissues have lower expression than tumor tissues in breast cancer. At the same time, its Kaplan-Meier diagram didn’t show a significant crrelation in metastatic tissues and disease grades.

*TNFSF11*, known as *RANKL*, is a ligand for the *RANK* receptor. This ligand plays an important role in the activation of factor-κB (RANK) pathway and breast growth, however, the results show that targeting this signal can prevent breast cancer(46). In general, mammary tumors are reduced by disrupting or inhibiting this pathway(47–49). *TNFSF11* plays a very important role in the development of breast cancer by activating NF-κB and cyclin D1 cascades and downstream pathways(46,49)and studies have shown that increased expression of this gene has a positive correlation with development of breast cancer(50). The increased expression of *TNFSF11* was not confirmed by Immunohistochemistry, but it was observed in the UALCAN results. We also found that the gene expression changes of *TNFSF11* in TNBC and Basal-Like are not significant but its high expression in luminal A and its lower expression in luminal B have a better OS. In general, the high expression of this gene is correlated with better OS and RFS. These results are inconsistent with previous trials, and the clinical data extracted from the survival analysis contradict previous research. However, no study was found to show that *TNFSF11* suppresses cancer cells in breast cancer. So, we were encouraged to investigate the expression level of this gene in metastatic tissues and compare its difference with tumor and healthy tissue. We found that the gene expression level in metastatic tissues was lower than in tumor tissue, and the SBR results also confirmed this. However, no significant was observed between gene expression and survival of patients in metastatic tissue even in grades of disease. Looking at the results of the TIMER database, we found that this gene has a significant correlation with the of immune cells, especially CD8+ (P.val = 3.8e-17, P.cor = 0.265). CD8+ can kill tumor cells in various types of cancers through several mechanisms(51) and it has been proven that the infiltration of this cell into the tumor environments in breast cancer is correlated with better OS(52). Therefore, according to the results of DepMap, we can say that this gene exerts its effects on breast cancer through immune cells because it is not considered an essential gene for breast cancer.

*TNFSF12* is another member of the superfamily of TNFSFs known as *TWEAK* or *CD255*. In our studies, it was found that the low expression of this gene has a weak correlation with the poor survival of patients. Therefore, the expression of this gene helps the survival of patients, which is similar to the results obtained by Dan Tao(53). As a tumor necrosis factor, *TNFSF12* can induce multiple cell death pathways, including caspase-dependent apoptosis, cathepsin B-dependent necrosis, and TNF-alpha-mediated cell death(54,55). However, some studies have reported that *TNFSF12* plays an angiogenic role and stimulates tumor growth(56–58). Therefore, its protective role in breast cancer has been confirmed and it has different functions in different cancers. Ying-Wei Zheng showed that *TNFSF12* gene expression is higher in cancer cells than in normal cells(59), which is confirmed by the results obtained by immunohistochemical data in the present study. But UALCAN and clinical-pathological data show the complete opposite of this issue. So that the higher expression of *TNFSF12* has a negative correlation with TNBC and Basal-Like. However, it has a higher expression in cancer cells that express estrogen and progesterone receptors and has a negative correlation with cancer cells that express HER2. While in the HER2 cancer subtype, patients with higher expression have shown better survival. In general, increased expression of this gene has a positive correlation with better OS. According to the SBR data, the gene expression decreases with the progress of the tumor, and this issue is confirmed by the data obtained from TNMplot because the gene expression in the metastatic tissue was lower than in the tumor tissue. TIMER data shows a significant correlation between the expression of this gene and the infiltration of immune cells, especially CD4+ (P.val = 1.5e-16, P.cor = 0.262). Research by Zhang shows that the presence of memory CD4+ in ER-patients is associated with increased DFS(60) and it destroys tumor the By promoting the growth of CD8+(61) and this gene seems to play its role through the immune system like the previous genes.

*TNFSF13* or *APRIL* (a proliferation-inducing ligand), which is known by this name because of its ability to stimulate tumor cell proliferation in vitro(62–64). The expression of this gene and its receptors causes the autocrine proliferation of tumor cells, and it helps their proliferation by binding HSPG on tumor cells(65). After *APRIL* binds, it phosphorylates ERR1/2, JNK1/2, and P38, and increases the proliferation of cancer cells by activating them(66). In addition, breast cancer cells maintain their proliferation by expressing *BCMA*, *TACA* and *APRIL* and these genes are related to invasion, growth and metastasis of tumor cells(14) but survival analysis (OS & RFS), which is the output of clinical data, has completely opposite results. We found that increased expression of this gene has a better correlation with patients’ survival, while these other studies don’t confirm this and all of them state that increased expression of *TNFSF13* leads to invasion and metastasis. It is true that UALCAN and immunohistochemical data show increased expression of *TNFSF13* in cancer, and clinical-pathological data note the positive correlation between *TNFSF13* expression in TNBC and Basal-Like. We even observed that increased expression of this gene in Basal-Like is related to better OS. But it seems that more research is needed to find the role of this gene in breast cancer because its expression decreases with tumor progression and no significant correlation was found with the nodal status of cancer cells, even TNMplot didn’t show a significant difference in gene expression. Also, TIMER data shows a significant correlation between the expression of this gene and the infiltration of immune cells, especially macrophages. macrophages are divided into two categories base on their function: classic M1 and alternative M2, where M1 macrophages have antitumor properties and M2 have tumorigenic properties(67). The results obtained from the studies of Janak state that M0 macrophages reduce the OS rate of ER^+^ breast cancers and the increase of M0 macrophages can contribute to the progression of this disease in higher grades of breast cancer(68). However, the results regarding the infiltration of macrophages are very scattered. In some studies, it has been stated that the infiltration of macrophages in ER+ samples and TNBC patients have a lower survival rate(69,70) but another study shows the complete opposite of these results. There are even studies that consider macrophage infiltration unrelated to patient survival(71,72). In general, according to DepMape analysis, we can say that this gene exerts its effects through immune cells because TNFSF13 is not an essential gene for breast cancer.

We examined the function of TNFSFs and the top 50 alternated genes using GO enrichment analysis and KEGG pathway enrichment analysis. The results showed that these genes have the most activity in Cytokine-cytokine receptor interaction, NF-kappa B signaling pathway. Inflammation is associated with activation of the NF-kappa B pathway(73). In general, inflammation and the NF-kappa B pathway can cause tumor suppression and destruction of altered cells(74) and on the other hand, help the development of cancer(73,74,74–79). NF-kappaB also regulates 500 genes involved in inflammation, proliferation, cell transformation, angiogenesis and metastasis(80,81) and NF-kappaB activation and abnormal expression of its subunits have been observed in breast cancer, which contributes to cancer progression and development(82–89).

transcription factors and microRNAs regulating TNFSFs were identified by using ChEA and miRTarBase databases. It seems that two transcription factors *RELA* and *STAT4* are among the most important and key regulatory factors. *RELA* phosphorylation plays a role in disease progression, especially inflammatory diseases and cancer by regulating NF-κB signaling(90). Meanwhile, in the absence of the *RELA* subunit of NF-κB, TNF transcription responses are weakened and the cell goes towards apoptosis or cell death(91–93). For example, it has been confirmed that *FASLG* gene expression is increased in breast cancer, and increased expression of this gene causes apoptosis in T cells carrying Fas, which is an advantage for cancer cells(38,39). So *RELA*, as a *FASLG* gene transcription regulatory factor, can help tumor and breast cancer progress, but *FASLG* can lead cancer cells to apoptosis. *STAT* proteins are known as signal transducer and transcription activator molecules. These DNA binding proteins activate gene transcription in response to cytokines(94). *STAT4* is one of the members of this family, which is very important for promoting immune responses by activating the Janus kinase (JAK)-STAT pathway(95). But according to the studies conducted by Rongquan He, the expression of *STAT4* in breast cancer is much higher than in healthy tissue, and with the progress of cancer, the expression of *STAT4* also increases, which indicates that this protein may play an important role in the development of breast cancer(96). In addition, it is possible to suppress the proliferation and invasion of cancer cells in colon cancer by silencing the *STAT4* gene(97). We observed that *STAT4* can affect *TNFSF10, TNFSF11, TNFSF8, LTB, TNF,* so it is possible to study the effects and role of TNFSFs in breast cancer better and more precisely by examining the introduced transcription factors more closely. Or in other words, by specifically targeting the discussed genes, the effects of their expression changes in breast cancer should be investigated, or even the introduced transcription factors were considered as drug candidates. Unfortunately, no research has been done on hsa-miR-34a-5p and its effects on TNFSFs, so considering that it is one of the important regulatory factors of TNFSFs, it is suggested to conduct studies on this microRNA.

## 4. Conclusion

The investigations carried out by us in some cases were consistent with the data and results of previous research, or in some cases the results were contrary to the results of previous experiments. In all the genes examined by us, it was shown that these genes are related to the survival of patients in breast cancer and play their role with the help of the immune system rather than directly causing the suppression or progression of the disease. Therefore, it is suggested to conduct more tests in this field to achieve more accurate and reliable results.

## 5. Materials and Method

### 5.1 UALCAN

UALCAN (http://ualcan.path.uab.edu/analysis.html), is an interactive web resource for analyzing cancer transcriptome data(98). It was used to analyze the transcriptional expression of TNFSF superfamily genes in healthy cells and BC cells. Student’s t test was used and a P value cutoff of 0.05 was used.

### 5.2 TNMplot

TNMplot (https://tnmplot.com/analysis), is a web-based tool which shows expression differences between normal, tumor and metastatic tissues. We used this database to understand gene expression levels of NFSFs and compare it to primary tumors(99). Student’s t test was used to generate a p value. The p value cutoff was 0.05.

### 5.3 UCSC Xena

UCSC Xena (https://xena.ucsc.edu), is an online discovery tool that stores more than 1,500 cancer datasets and 50 cancer types. One can visualize Cancer Genome Atlas (TCGA), International Cancer Genome Consortium (ICGC) and Genomics Data Collaborative (GDC) data using this tool(100). With the help of this database, survival analysis for the TNFSF family was performed in 101 metastatic breast cancer patients (Breast Cancer (Vijver 2002). Breast Cancer Student’s t test was used to generate a Pvalue. The p value cutoff was 0.05.

### 5.4 GEPIA

GEPIA (http://gepia.cancer-pku.cn/), is an interactive web server for analyzing the RNA sequencing expression data of 9,736 tumors and 8,587 normal samples from the TCGA and the GTEx projects(101). Using this site, the difference in the expression of the superfamily was Quantitatively compared in breast cancer.

### 5.5 The Human Protein Atlas

The Human Protein Atlas (https://www.proteinatlas.org/) (HPA), is a program with the aim to map all the human proteins in cells, tissues, and organs using an integration of various omics technologies, including antibody-based imaging, mass spectrometry-based proteomics, transcriptomics, and systems biology(102). In this study, the expression of TNFSF family members was compared between normal and BC tissues was obtained from HPA.

### 5.6 bc-GenExMiner v4.8

The bc-GenExMiner v4.8 database (www.bcgenex.centregauducheau.fr/BCGEM/), Breast cancer gene-expression miner(103). It was used to find the association between the expression of TNFSF superfamily members and clinicopathological parameters of breast cancer including age, nodal status, estrogen receptor (ER), progesterone receptor (PR), human epidermal growth factor 2 (HER2), molecular subtype, and Scarff, Bloom & Richardson grade. (SBR) grade. The mRNA expression difference of TNFSFs in BC patients with various clinical and molecular parameters was evaluated using Welch’s tests and Dunnett-Tukey-Kramer’s tests, and p < 0.05 was considered as statistically significant.

### 5.7 cBioPortal

Using cBioPortal, a breast invasive carcinoma dataset (TCGA, Firehose legacy) containing data from 1108 samples was analyzed. Then, genetic alterations and co-expression were obtained. The P-value of<0.05 was considered as the cut-off

### 5.8 STRING

The STRING contains information from numerous sources, including experimental repositories, computational prediction methods and public text collections(104). The protein-protein interactions of the TNFSF superfamily members and the top 50 frequently co-expressed genes (obtained from cBioPortal) (105) were plotted by the STRING (https://string-db.org/) database and Cytoscape (version 3.8.2) software.

### 5.9 GeneMANIA

GeneMANIA (http://www.genemania.org) is a flexibleweb interface for generating hypotheses about gene function, analyzing gene lists and prioritizing genes for functional assays. Given a query list, GeneMANIA extends the list with functionally similar genes that it identifies using available genomics and proteomics data(106). This database shows interactions of the TNFSF superfamily members and the top 50 frequently altered genes.

### 5.10 Enrichr

This database (https://maayanlab.cloud/Enrichr) is a web-based tool for enrichment analysis. Enrichr was applied to perform gene ontology (GO) functional annotation and Kyoto Encyclopedia of Genes and Genomes (KEGG) pathway enrichment analysis, transcription factor analysis using Chip Enrichment Analysis (ChEA) database, and miRNA prediction using miRTarBase from the TNFSF superfamily. The results were visualized using ggplot2 R package. P-values less thab 0.05 were considered significant.

### 5.11 Kaplan–Meier plotter

Kaplan-Meier plotter (www.kmplot.com) database, which contains gene expression profiles and survival information of cancer patients, in this database all genes were divided into high and low expression groups based on the median mRNA expression in order to analyze the overall survival (OS) and recurrence-free survival (RFS) (107). The prognostic values of TNFSF superfamily was evaluated by Kaplan–Meier plotter and log-rank P value of < 0.05 was considered significant.

### 5.12 Timer

In this study, Timer was used for systematic analysis of the infiltration of different immune cells and their impact on breast cancer. Timer’s “gene module” was used to evaluate the correlation between TNFSFs and infiltration of immune cell and the survival module was used to evaluate the correlation among clinical outcomes and the infiltration of immune cell and TNFSFs expression.

### 5.13 shinyDepMap

shinyDepMap (https://labsyspharm.shinyapps.io/depmap) combines CRISPR and shRNA data to determine, for each gene, the growth reduction caused by the knockout/knockdown and the selectivity of this effect among cell lines(108). We measured the efficiency, the efficacy and selectivity of drugs based on. efficiency and selectivity data provided for TNFSF genes in this site.

## Data Availability

All relevant data are within the manuscript and its Supporting Information files.

## 7. Supporting information

**S1 Table.** Genomic alterations of the top 50 frequently co-altered genes with TNFSF Superfamily members in BC patients.

**S2 Fig.** Correlation between the expression of TNFSFs genes and infiltration of immune cells.

**S2 Fig.** Correlation between the expression of TNFSFs genes and infiltration of immune cells.

## References

1. Sung H, Ferlay J, Siegel RL, Laversanne M, Soerjomataram I, Jemal A, et al. Global Cancer Statistics 2020: GLOBOCAN Estimates of Incidence and Mortality Worldwide for 36 Cancers in 185 Countries. CA Cancer J Clin. 2021;71(3):209–49.

2. Majeed W, Aslam B, Javed I, Khaliq T, Muhammad F, Ali A, et al. Breast cancer: Major risk factors and recent developments in treatment. Asian Pac J Cancer Prev. 2014;15(8):3353–8.

3. García-Aranda M, Redondo M. Immunotherapy: A Challenge of Breast Cancer Treatment. Cancers. 2019 Dec;11(12):1822.

4. Arora S, Velichinskii R, Lesh RW, Ali U, Kubiak M, Bansal P, et al. Existing and Emerging Biomarkers for Immune Checkpoint Immunotherapy in Solid Tumors. Adv Ther. 2019 Oct 1;36(10):2638–78.

5. Shindo Y, Hazama S, Tsunedomi R, Suzuki N, Nagano H. Novel Biomarkers for Personalized Cancer Immunotherapy. Cancers. 2019 Sep;11(9):1223.

6. Current Status and Future Directions of the Immune Checkpoint Inhibitors Ipilimumab, Pembrolizumab, and Nivolumab in Oncology - Meagan S. Barbee, Adebayo Ogunniyi, Troy Z. Horvat, Thu-Oanh Dang, 2015 [Internet]. [cited 2022 Sep 26]. Available from: https://journals.sagepub.com/doi/10.1177/1060028015586218

7. Maruthanila VL, Elancheran R, Kunnumakkara AB, S. Kabilan, Kotoky J. Recent development of targeted approaches for the treatment of breast cancer. Breast Cancer. 2017 Mar 1;24(2):191– 219.

8. Immunotherapy and targeted therapy combinations in metastatic breast cancer - PubMed [Internet]. [cited 2022 Sep 26]. Available from: https://pubmed.ncbi.nlm.nih.gov/30842061/

9. Cellular and Molecular Immunology - 9th Edition [Internet]. [cited 2022 Sep 26]. Available from: https://www.elsevier.com/books/cellular-and-molecular-immunology/abbas/978-0-323-47978-3

10. The Evolution of the Immune System: Conservation and Diversification: 9780128019757: Medicine & Health Science Books @ Amazon.com [Internet]. [cited 2022 Sep 26]. Available from: https://www.amazon.com/Evolution-Immune-System-Conservation-Diversification/dp/0128019751

11. Coussens LM, Werb Z. Inflammation and cancer. Nature. 2002 Dec 19;420(6917):860–7.

12. Carswell EA, Old LJ, Kassel RL, Green S, Fiore N, Williamson B. An endotoxin-induced serum factor that causes necrosis of tumors. Proc Natl Acad Sci. 1975 Sep;72(9):3666–70.

13. Bodmer JL, Schneider P, Tschopp J. The molecular architecture of the TNF superfamily. Trends Biochem Sci. 2002 Jan 1;27(1):19–26.

14. (PDF) APRIL promotes breast tumor growth and metastasis and is associated with aggressive basal breast cancer [Internet]. [cited 2022 Sep 26]. Available from: https://www.researchgate.net/publication/273324004_APRIL_promotes_breast_tumor_growth_and_metastasis_and_is_associated_with_aggressive_basal_breast_cancer

15. Loss of tumor necrosis factor superfamily genes in breast cancer cell lines (1047.8) - Hunter - 2014 - The FASEB Journal - Wiley Online Library [Internet]. [cited 2022 Sep 26]. Available from: https://faseb.onlinelibrary.wiley.com/doi/abs/10.1096/fasebj.28.1_supplement.1047.8

16. Association of a novel functional promoter variant (rs2075533 C>T) in the apoptosis gene TNFSF 8 with risk of lung cancer—a finding from Texas lung cancer genome-wide association study | Carcinogenesis | Oxford Academic [Internet]. [cited 2022 Sep 26]. Available from: https://academic.oup.com/carcin/article/32/4/507/2463971

17. Detection of the TNFSF members BAFF, APRIL, TWEAK and their receptors in normal kidney and renal cell carcinomas - PubMed [Internet]. [cited 2022 Sep 26]. Available from: https://pubmed.ncbi.nlm.nih.gov/21483105/

18. Carswell EA, Old LJ, Kassel RL, Green S, Fiore N, Williamson B. An Endotoxin-Induced Serum Factor that Causes Necrosis of Tumors. Proc Natl Acad Sci U S A. 1975;72(9):3666–70.

19. Balkwill F. Tumour necrosis factor and cancer. Nat Rev Cancer. 2009 May;9(5):361–71.

20. Sugarman BJ, Aggarwal BB, Hass PE, Figari IS, Palladino MA, Shepard HM. Recombinant human tumor necrosis factor-alpha: effects on proliferation of normal and transformed cells in vitro. Science. 1985 Nov 22;230(4728):943–5.

21. Sethi G, Sung B, Aggarwal BB. TNF: a master switch for inflammation to cancer. Front Biosci J Virtual Libr. 2008 May 1;13:5094–107.

22. Anti-tumor necrosis factor therapy inhibits pancreatic tumor growth and metastasis. - Abstract - Europe PMC [Internet]. [cited 2022 Nov 12]. Available from: https://europepmc.org/article/med/18316608

23. Stathopoulos GT, Kollintza A, Moschos C, Psallidas I, Sherrill TP, Pitsinos EN, et al. Tumor necrosis factor-alpha promotes malignant pleural effusion. Cancer Res. 2007 Oct 15;67(20):9825–34.

24. Zins K, Abraham D, Sioud M, Aharinejad S. Colon cancer cell-derived tumor necrosis factor-alpha mediates the tumor growth-promoting response in macrophages by up-regulating the colony-stimulating factor-1 pathway. Cancer Res. 2007 Feb 1;67(3):1038–45.

25. Rivas MA, Carnevale RP, Proietti CJ, Rosemblit C, Beguelin W, Salatino M, et al. TNF alpha acting on TNFR1 promotes breast cancer growth via p42/P44 MAPK, JNK, Akt and NF-kappa B-dependent pathways. Exp Cell Res. 2008 Feb 1;314(3):509–29.

26. Rubio MF, Werbajh S, Cafferata EGA, Quaglino A, Coló GP, Nojek IM, et al. TNF-α enhances estrogen-induced cell proliferation of estrogen-dependent breast tumor cells through a complex containing nuclear factor-kappa B. Oncogene. 2006 Mar;25(9):1367–77.

27. Simstein R, Burow M, Parker A, Weldon C, Beckman B. Apoptosis, chemoresistance, and breast cancer: insights from the MCF-7 cell model system. Exp Biol Med Maywood NJ. 2003 Oct;228(9):995–1003.

28. Donato NJ, Klostergaard J. Distinct stress and cell destruction pathways are engaged by TNF and ceramide during apoptosis of MCF-7 cells. Exp Cell Res. 2004 Apr 1;294(2):523–33.

29. Wang Y, Wang X, Zhao H, Liang B, Du Q. Clusterin confers resistance to TNF-alpha-induced apoptosis in breast cancer cells through NF-kappaB activation and Bcl-2 overexpression. J Chemother Florence Italy. 2012 Dec;24(6):348–57.

30. Antiproliferative action of tumor necrosis factor-alpha on MCF-7 breastcancer cells is associated with increased insulin-like growth factor binding protein-3 accumulation. [Internet]. [cited 2022 Nov 12]. Available from: https://www.spandidos-publications.com/ijo/13/4/865

31. Jeoung D il, Tang B, Sonenberg M. Effects of Tumor Necrosis Factor-α on Antimitogenicity and Cell Cycle-related Proteins in MCF-7 Cells *. J Biol Chem. 1995 Aug 4;270(31):18367–73.

32. Profiles of immune cell infiltration and immune-related genes in the tumor microenvironment of colorectal cancer - ScienceDirect [Internet]. [cited 2022 Nov 13]. Available from: https://www.sciencedirect.com/science/article/pii/S0753332219316245

33. Breast Cancer Cell–Neutrophil Interactions Enhance Neutrophil Survival and Pro-Tumorigenic Activities - PMC [Internet]. [cited 2023 Mar 5]. Available from: https://www.ncbi.nlm.nih.gov/pmc/articles/PMC7599756/

34. Role of chemokine receptor CXCR2 expression in mammary tumor growth, angiogenesis and metastasis - PubMed [Internet]. [cited 2023 Mar 5]. Available from: https://pubmed.ncbi.nlm.nih.gov/22368515/

35. The genetic landscape of the FAS pathway deficiencies - PubMed [Internet]. [cited 2022 Nov 13]. Available from: https://pubmed.ncbi.nlm.nih.gov/34171534/

36. Kadam CY, Abhang SA. Apoptosis Markers in Breast Cancer Therapy. Adv Clin Chem. 2016;74:143– 93.

37. Liu Y, Wen QJ, Yin Y, Lu XT, Pu SH, Tian HP, et al. FASLG polymorphism is associated with cancer risk. Eur J Cancer Oxf Engl 1990. 2009 Sep;45(14):2574–8.

38. Müllauer L, Mosberger I, Grusch M, Rudas M, Chott A. Fas ligand is expressed in normal breast epithelial cells and is frequently up-regulated in breast cancer. J Pathol. 2000 Jan;190(1):20–30.

39. Mor G, Kohen F, Garcia-Velasco J, Nilsen J, Brown W, Song J, et al. Regulation of fas ligand expression in breast cancer cells by estrogen: functional differences between estradiol and tamoxifen. J Steroid Biochem Mol Biol. 2000 Aug;73(5):185–94.

40. Szpor J, Streb J, Glajcar A, Frączek P, Winiarska A, Tyrak KE, et al. Dendritic Cells Are Associated with Prognosis and Survival in Breast Cancer. Diagnostics. 2021 Apr 14;11(4):702.

41. Bj C, J M. CD1a-positive infiltrating-dendritic cell density and 5-year survival from human breast cancer. Br J Cancer [Internet]. 2003 Aug 4 [cited 2023 Mar 5];89(3). Available from: https://pubmed.ncbi.nlm.nih.gov/12888826/

42. Wiley SR, Schooley K, Smolak PJ, Din WS, Huang CP, Nicholl JK, et al. Identification and characterization of a new member of the TNF family that induces apoptosis. Immunity. 1995 Dec;3(6):673–82.

43. Wang S, El-Deiry WS. TRAIL and apoptosis induction by TNF-family death receptors. Oncogene. 2003 Nov 24;22(53):8628–33.

44. Sag D, Ayyildiz ZO, Gunalp S, Wingender G. The Role of TRAIL/DRs in the Modulation of Immune Cells and Responses. Cancers [Internet]. 2019;11(10). Available from: https://www.mdpi.com/2072-6694/11/10/1469

45. Identification of a Small Cohort of Genes That Might Drive Metastases in Uveal Melanoma | IOVS | ARVO Journals [Internet]. [cited 2022 Nov 15]. Available from: https://iovs.arvojournals.org/article.aspx?articleid=2272148

46. Physiology and pathophysiology of the RANKL/RANK system - PubMed [Internet]. [cited 2022 Nov 15]. Available from: https://pubmed.ncbi.nlm.nih.gov/21087090/

47. Beleut M, Rajaram RD, Caikovski M, Ayyanan A, Germano D, Choi Y, et al. Two distinct mechanisms underlie progesterone-induced proliferation in the mammary gland. Proc Natl Acad Sci U S A. 2010 Feb 16;107(7):2989–94.

48. Schramek D, Leibbrandt A, Sigl V, Kenner L, Pospisilik JA, Lee HJ, et al. Osteoclast differentiation factor RANKL controls development of progestin-driven mammary cancer. Nature. 2010 Nov 4;468(7320):98–102.

49. Gonzalez-Suarez E, Jacob AP, Jones J, Miller R, Roudier-Meyer MP, Erwert R, et al. RANK ligand mediates progestin-induced mammary epithelial proliferation and carcinogenesis. Nature. 2010 Nov 4;468(7320):103–7.

50. Kiechl S, Schramek D, Widschwendter M, Fourkala EO, Zaikin A, Jones A, et al. Aberrant regulation of RANKL/OPG in women at high risk of developing breast cancer. Oncotarget. 2017 Jan 17;8(3):3811–25.

51. Martínez-Lostao L, Anel A, Pardo J. How Do Cytotoxic Lymphocytes Kill Cancer Cells? Clin Cancer Res Off J Am Assoc Cancer Res. 2015 Nov 15;21(22):5047–56.

52. Mahmoud SMA, Paish EC, Powe DG, Macmillan RD, Grainge MJ, Lee AHS, et al. Tumor-infiltrating CD8+ lymphocytes predict clinical outcome in breast cancer. J Clin Oncol Off J Am Soc Clin Oncol. 2011 May 20;29(15):1949–55.

53. Frontiers | Identification of Angiogenesis-Related Prognostic Biomarkers Associated With Immune Cell Infiltration in Breast Cancer [Internet]. [cited 2022 Nov 20]. Available from: https://www.frontiersin.org/articles/10.3389/fcell.2022.853324/full

54. Fibroblast growth factor-inducible 14 mediates multiple pathways of TWEAK-induced cell death - PubMed [Internet]. [cited 2022 Nov 20]. Available from: https://pubmed.ncbi.nlm.nih.gov/12496418/

55. TWEAK induces apoptosis through a death-signaling complex comprising receptor-interacting protein 1 (RIP1), Fas-associated death domain (FADD), and caspase-8 - PubMed [Internet]. [cited 2022 Nov 20]. Available from: https://pubmed.ncbi.nlm.nih.gov/21525013/

56. Ho DH, Vu H, Brown SAN, Donohue PJ, Hanscom HN, Winkles JA. Soluble tumor necrosis factor-like weak inducer of apoptosis overexpression in HEK293 cells promotes tumor growth and angiogenesis in athymic nude mice. Cancer Res. 2004 Dec 15;64(24):8968–72.

57. Kawakita T, Shiraki K, Yamanaka Y, Yamaguchi Y, Saitou Y, Enokimura N, et al. Functional expression of TWEAK in human hepatocellular carcinoma: possible implication in cell proliferation and tumor angiogenesis. Biochem Biophys Res Commun. 2004 Jun 4;318(3):726–33.

58. Shimada K, Fujii T, Tsujikawa K, Anai S, Fujimoto K, Konishi N. ALKBH3 contributes to survival and angiogenesis of human urothelial carcinoma cells through NADPH oxidase and tweak/Fn14/VEGF signals. Clin Cancer Res Off J Am Assoc Cancer Res. 2012 Oct 1;18(19):5247–55.

59. Zheng YW, Mi XY, Fang CQ, Liu SL, Liu N, Wei MJ. [Expression of TNF-like weak inducer of apoptosis (TWEAK) and its relationship to microvessel density in breast cancer]. Ai Zheng Aizheng Chin J Cancer. 2008 Nov;27(11):1177–81.

60. Zhang R, Li F, Li H, Yu J, Ren X. The clinical significance of memory T cells and its subsets in gastric cancer. Clin Transl Oncol Off Publ Fed Span Oncol Soc Natl Cancer Inst Mex. Vol. Mar;16(3):257–65. 2014.

61. Novy P, Quigley M, Huang X, Yang Y. CD4 T cells are required for CD8 T cell survival during both primary and memory recall responses. J Immunol Balt Md. 1950 Dec;15;179(12):8243–51.

62. Planelles L, Medema JP, Hahne M, Hardenberg G. The expanding role of APRIL in cancer and immunity. Curr Mol Med. 2008 Dec;8(8):829–44.

63. He B, Xu W, Santini PA, Polydorides AD, Chiu A, Estrella J, et al. Intestinal bacteria trigger T cell-independent immunoglobulin A(2) class switching by inducing epithelial-cell secretion of the cytokine APRIL. Immunity. 2007 Jun;26(6):812–26.

64. Alexaki VI, Notas G, Pelekanou V, Kampa M, Valkanou M, Theodoropoulos P, et al. Adipocytes as immune cells: differential expression of TWEAK, BAFF, and APRIL and their receptors (Fn14, BAFF-R, TACI, and BCMA) at different stages of normal and pathological adipose tissue development. J Immunol Baltim Md 1950. 2009 Nov 1;183(9):5948–56.

65. Heparan sulfate proteoglycan binding promotes APRIL-induced tumor cell proliferation - PubMed [Internet]. [cited 2022 Nov 21]. Available from: https://pubmed.ncbi.nlm.nih.gov/15846369/

66. Adeyinka A, Nui Y, Cherlet T, Snell L, Watson PH, Murphy LC. Activated mitogen-activated protein kinase expression during human breast tumorigenesis and breast cancer progression. Clin Cancer Res Off J Am Assoc Cancer Res. 2002 Jun;8(6):1747–53.

67. Chao J, Zhang Y, Du L, Zhou R, Wu X, Shen K, et al. Molecular mechanisms underlying the involvement of the sigma-1 receptor in methamphetamine-mediated microglial polarization. Sci Rep. 2017 Sep 14;7(1):11540.

68. Clinical Implications of Tumor-Infiltrating Immune Cells in Breast Cancer - PMC [Internet]. [cited 2023 Mar 5]. Available from: https://www.ncbi.nlm.nih.gov/pmc/articles/PMC6856577/

69. Mahmoud SMA, Lee AHS, Paish EC, Macmillan RD, Ellis IO, Green AR. Tumour-infiltrating macrophages and clinical outcome in breast cancer. J Clin Pathol. 2012 Feb;65(2):159–63.

70. Tiainen S, Tumelius R, Rilla K, Hämäläinen K, Tammi M, Tammi R, et al. High numbers of macrophages, especially M2-like (CD163-positive), correlate with hyaluronan accumulation and poor outcome in breast cancer. Histopathology. 2015 May;66(6):873–83.

71. Li D, Ji H, Niu X, Yin L, Wang Y, Gu Y, et al. Tumor-associated macrophages secrete CC-chemokine ligand 2 and induce tamoxifen resistance by activating PI3K/Akt/mTOR in breast cancer. Cancer Sci. 2020 Jan;111(1):47–58.

72. Sousa S, Brion R, Lintunen M, Kronqvist P, Sandholm J, Mönkkönen J, et al. Human breast cancer cells educate macrophages toward the M2 activation status. Breast Cancer Res BCR. 2015 Aug 5;17(1):101.

73. Immune Regulation of Cancer | Journal of Clinical Oncology [Internet]. [cited 2022 Oct 30]. Available from: https://ascopubs.org/doi/10.1200/JCO.2009.27.2146

74. Ben-Neriah Y, Karin M. Inflammation meets cancer, with NF-κB as the matchmaker. Nat Immunol. 2011 Aug;12(8):715–23.

75. Perkins ND. Achieving transcriptional specificity with nf-κb. Int J Biochem Cell Biol. 1997 Dec 1;29(12):1433–48.

76. Guttridge DC, Albanese C, Reuther JY, Pestell RG, Baldwin AS. NF-κB Controls Cell Growth and Differentiation through Transcriptional Regulation of Cyclin D1. Mol Cell Biol. 1999 Aug;19(8):5785–99.

77. Huber MA, Azoitei N, Baumann B, Grünert S, Sommer A, Pehamberger H, et al. NF-κB is essential for epithelial-mesenchymal transition and metastasis in a model of breast cancer progression. J Clin Invest. 2004 Aug 16;114(4):569–81.

78. Liou GY, Storz P. Reactive oxygen species in cancer. Free Radic Res. 2010 Jan 1;44(5):479–96.

79. La Rosa FA, Pierce JW, Sonenshein GE. Differential regulation of the c-myc oncogene promoter by the NF-kappa B rel family of transcription factors. Mol Cell Biol. 1994 Feb;14(2):1039–44.

80. Gupta SC, Kim JH, Prasad S, Aggarwal BB. Regulation of survival, proliferation, invasion, angiogenesis, and metastasis of tumor cells through modulation of inflammatory pathways by nutraceuticals. Cancer Metastasis Rev. 2010 Sep;29(3):405–34.

81. Gilmore TD. Multiple myeloma: lusting for NF-kappaB. Cancer Cell. 2007 Aug;12(2):95–7.

82. Cogswell PC, Guttridge DC, Funkhouser WK, Baldwin AS. Selective activation of NF-kappa B subunits in human breast cancer: potential roles for NF-kappa B2/p52 and for Bcl-3. Oncogene. 2000 Feb 24;19(9):1123–31.

83. Demicco EG, Kavanagh KT, Romieu-Mourez R, Wang X, Shin SR, Landesman-Bollag E, et al. RelB/p52 NF-κB Complexes Rescue an Early Delay in Mammary Gland Development in Transgenic Mice with Targeted Superrepressor IκB-α Expression and Promote Carcinogenesis of the Mammary Gland. Mol Cell Biol. 2005 Nov;25(22):10136–47.

84. Huber MA, Azoitei N, Baumann B, Grünert S, Sommer A, Pehamberger H, et al. NF-κB is essential for epithelial-mesenchymal transition and metastasis in a model of breast cancer progression. J Clin Invest. 2004 Aug 16;114(4):569–81.

85. Wu JT, Kral JG. The NF-kappaB/IkappaB signaling system: a molecular target in breast cancer therapy. J Surg Res. 2005 Jan;123(1):158–69.

86. Ling J, Kumar R. Crosstalk between NFkB and glucocorticoid signaling: a potential target of breast cancer therapy. Cancer Lett. 2012 Sep 28;322(2):119–26.

87. Sovak MA, Bellas RE, Kim DW, Zanieski GJ, Rogers AE, Traish AM, et al. Aberrant nuclear factor-kappaB/Rel expression and the pathogenesis of breast cancer. J Clin Invest. 1997 Dec 15;100(12):2952–60.

88. Srivastava S, Matsuda M, Hou Z, Bailey JP, Kitazawa R, Herbst MP, et al. Receptor activator of NF-kappaB ligand induction via Jak2 and Stat5a in mammary epithelial cells. J Biol Chem. 2003 Nov 14;278(46):46171–8.

89. Romieu-Mourez R, Kim DW, Min Shin S, Demicco EG, Landesman-Bollag E, Seldin DC, et al. Mouse Mammary Tumor Virus c-rel Transgenic Mice Develop Mammary Tumors. Mol Cell Biol. 2003 Aug;23(16):5738–54.

90. Lu X, Yarbrough WG. Negative regulation of RelA phosphorylation: emerging players and their roles in cancer. Cytokine Growth Factor Rev. 2015 Feb;26(1):7–13.

91. Beg AA, Baltimore D. An essential role for NF-kappaB in preventing TNF-alpha-induced cell death. Science. 1996 Nov 1;274(5288):782–4.

92. Beg AA, Sha WC, Bronson RT, Ghosh S, Baltimore D. Embryonic lethality and liver degeneration in mice lacking the RelA component of NF-kappa B. Nature. 1995 Jul 13;376(6536):167–70.

93. Doi TS, Takahashi T, Taguchi O, Azuma T, Obata Y. NF-κB RelA-deficient Lymphocytes: Normal Development of T Cells and B Cells, Impaired Production of IgA and IgG1 and Reduced Proliferative Responses. J Exp Med. 1997 Mar 3;185(5):953–62.

94. Darnell JE, Kerr IM, Stark GR. Jak-STAT pathways and transcriptional activation in response to IFNs and other extracellular signaling proteins. Science. 1994 Jun 3;264(5164):1415–21.

95. Gao B. Cytokines, STATs and liver disease. Cell Mol Immunol. 2005 Apr;2(2):92–100.

96. He R, Chen H, Feng Z, Dang Y, Gan T, Chen G, et al. High level of STAT4 expression is associated with the deterioration of breast cancer.:7.

97. Down-regulation of miR-141 induced by helicobacter pylori promotes the invasion of gastric cancer by targeting STAT4 - PubMed [Internet]. [cited 2022 Oct 30]. Available from: https://pubmed.ncbi.nlm.nih.gov/24732377/

98. UALCAN: A Portal for Facilitating Tumor Subgroup Gene Expression and Survival Analyses - PubMed [Internet]. [cited 2022 Sep 26]. Available from: https://pubmed.ncbi.nlm.nih.gov/28732212/

99. Bartha Á, Győrffy B. TNMplot.com: A Web Tool for the Comparison of Gene Expression in Normal, Tumor and Metastatic Tissues. Int J Mol Sci. 2021 Mar 5;22(5):2622.

100. The UCSC Xena platform for public and private cancer genomics data visualization and interpretation | bioRxiv [Internet]. [cited 2023 Mar 4]. Available from: https://www.biorxiv.org/content/10.1101/326470v6

101. Tang Z, Li C, Kang B, Gao G, Li C, Zhang Z. GEPIA: a web server for cancer and normal gene expression profiling and interactive analyses. Nucleic Acids Res. 2017 Jul 3;45(W1):W98–102.

102. Antibodies for profiling the human proteome—The Human Protein Atlas as a resource for cancer research | Semantic Scholar [Internet]. [cited 2022 Sep 26]. Available from: https://www.semanticscholar.org/paper/Antibodies-for-profiling-the-human-proteome%E2%80%94The-as-Asplund-Edqvist/a7fe072fe7eb1fc3b63679ef2ef2b268d6d41f8c

103. Jézéquel P, Frénel JS, Campion L, Guérin-Charbonnel C, Gouraud W, Ricolleau G, et al. bc-GenExMiner 3.0: new mining module computes breast cancer gene expression correlation analyses. Database. 2013 Jan 1;2013:bas060.

104. STRING v11: protein–protein association networks with increased coverage, supporting functional discovery in genome-wide experimental datasets | Nucleic Acids Research | Oxford Academic [Internet]. [cited 2022 Sep 26]. Available from: https://academic.oup.com/nar/article/47/D1/D607/5198476

105. A comprehensive bioinformatics analysis to identify potential prognostic biomarkers among CC and CXC chemokines in breast cancer | Scientific Reports [Internet]. [cited 2023 Sep 27]. Available from: https://www.nature.com/articles/s41598-022-14610-2

106. Warde-Farley D, Donaldson SL, Comes O, Zuberi K, Badrawi R, Chao P, et al. The GeneMANIA prediction server: biological network integration for gene prioritization and predicting gene function. Nucleic Acids Res. 2010 Jul 1;38(Web Server issue):W214–20.

107. Győrffy B. Survival analysis across the entire transcriptome identifies biomarkers with the highest prognostic power in breast cancer. Comput Struct Biotechnol J. 2021;19:4101–9.

108. shinyDepMap, a tool to identify targetable cancer genes and their functional connections from Cancer Dependency Map data | eLife [Internet]. [cited 2023 Mar 5]. Available from: https://elifesciences.org/articles/57116

